# Temporal EHR Models Detect Rare Disease Years Before Diagnosis

**DOI:** 10.64898/2026.09.10.26362561

**Authors:** Lu Yang, Pauline Ng, Ming Yin Lun, Tate Tunstall, Lindsay Meyers, Mayowa Osundiji, Kate Im, Akash Kumar, Adam Lavertu, Matthew Rabinowitz

## Abstract

**Background:** Rare diseases collectively affect an estimated 300 million people worldwide, yet patients wait an average of 4–8 years for a correct diagnosis after visiting multiple physicians, accumulating unexplained findings, and suffering preventable disease progression before a unifying diagnosis is reached. Electronic health records (EHRs) capture this pre-diagnostic trajectory in rich detail, but conventional methods typically collapse years of longitudinal phenotypic signals into a single static snapshot; our approach instead models this temporal structure directly.

**Methods:** We conducted a retrospective cohort study using longitudinal EHR data from approximately 3 million patients at Mayo Clinic Platform_Accelerate. Cases were patients with confirmed diagnoses of 10 rare diseases; controls (∼5,200 per disease) had no record of any rare disease. HPO phenotype terms were extracted from clinical notes and laboratory results using a medspaCy NLP pipeline; ICD codes were grouped at three-character level. We developed and trained two temporal models, GRU-Attn and Conformer, that encode patient histories as quarterly time-binned sequences spanning up to 30 years, and compared these against three static baselines (CatBoost, XGBoost, logistic regression). Models were evaluated systematically at prediction horizons h = 1–10 years before diagnosis.

**Results:** In the full held-out cohort benchmark, temporal models outperformed static baselines across the large majority of diseases and prediction horizons, with the advantage widening substantially as the horizon lengthened. At 1–2 years prior to diagnosis, mean AUROC was 0.928 (GRU-Attn) versus 0.871 (CatBoost); at 6–10 years prior to diagnosis, 0.726 versus 0.693, with static models degrading to near-chance for some diseases. Combining HPO and ICD features yielded the strongest performance, with a mean gain of +0.063 AUROC over the best single source at 1–2 years prior to diagnosis. Separately, in a manually curated cohort of 143 confirmed-undiagnosed cases with extended EHR histories, GRU-Attn identified 138 cases before first disease mention and all 143 cases before formal diagnosis at the prespecified 99th-percentile specificity threshold, with median lead times of 1.1–8.3 years and 1.9–21.7 years, respectively.

**Conclusions:** By learning from the temporal trajectory of phenotypic signals rather than their static aggregate, temporal EHR models improved retrospective early detection performance across several rare diseases, most powerfully for conditions where pre-diagnostic findings accrue gradually and heterogeneously over years. These results show that longitudinal EHR trajectories carry rare-disease signal beyond static phenotypic burden and support prospective evaluation of temporal EHR modeling for rare-disease risk stratification.

## INTRODUCTION

Rare diseases affect an estimated 300 million people worldwide, yet individual conditions are frequently unrecognized or misdiagnosed for years; average diagnostic delays of 4–8 years are consistently reported^1–3^. For example, a patient with Fabry disease may visit rheumatologists, gastroenterologists, and dermatologists for years, with pain in the hands and feet, gastrointestinal symptoms, and angiokeratomas managed in isolation rather than recognized as a single disease. Similarly, a patient with hereditary angioedema may experience recurrent episodes of swelling and abdominal pain for years before the longitudinal pattern is recognized and a diagnosis reached. These delays carry profound consequences: disease progression during the diagnostic odyssey leads to irreversible organ damage, reduced treatment efficacy, and substantial psychological and financial burden for patients and families^4^. For conditions such as hereditary angioedema, where long-term prophylactic therapies can prevent life-threatening attacks, or Fabry disease, where enzyme replacement therapy can prevent progressive renal and cardiac damage, earlier diagnosis may enable earlier risk mitigation or surveillance in selected conditions.

EHRs contain the longitudinal phenotypic signals that precede formal diagnosis: patterns of specialist referrals, incidental laboratory findings, and symptom descriptions accumulate in the clinical record years before a unifying diagnosis is established^5–8^. Existing approaches, however, typically collapse these histories into static feature vectors, presence or count of each finding across all available time, discarding the temporal ordering that may be the most diagnostically informative dimension of the data. For example, unusually tall stature flagged at a paediatric visit at 10, a lens dislocation recorded by an ophthalmologist at 16, and an incidental note of aortic root widening on an echocardiogram at 24 may each seem unrelated; together they describe a Marfan syndrome. Or, a patient with an emergency visit for unexplained facial swelling at 13, a hospitalisation for severe abdominal pain with no clear cause at 19, and a subsequent swelling episode may each be coded as isolated events; together they suggest hereditary angioedema. Hospital visits and their symptoms may each be unremarkable in isolation; their temporal sequence and rate of accrual may be pathognomonic.

Several temporal deep learning architectures have been applied to common disease prediction in EHR data, including recurrent networks, transformers, and graph-based models^10–12^. However, rare diseases present a distinct challenge: small case counts, heterogeneous presentations, and decade-long pre-diagnostic trajectories demand models that can leverage sparse signals over extended time windows. No prior work has systematically evaluated temporal sequence models^13–15^, across a panel of rare diseases at multi-year prediction horizons or characterized the practical early detection lead times achievable at high-specificity thresholds intended to approximate potential screening settings. We further develop two novel architectures adapted for the rare disease problem: GRU-Attn, which extends recurrent modeling with 3-way attention multi-pooling to capture trajectory timing, average burden, and transient early signals simultaneously, and Conformer-Attn, which combines local convolutional and global self-attention mechanisms to capture both short-range phenotypic clusters and long-range temporal dependencies across decade-long EHR histories.

We present a systematic evaluation of temporal sequence models, GRU-Attn and Conformer, for early detection of 10 rare diseases across prediction horizons of 1-10 years before diagnosis. We compare these against static baselines and characterize potentially actionable lead times at high-specificity thresholds, supporting prospective evaluation rather than immediate deployment.

## METHODS

### Study Design and Cohort

We conducted a retrospective cohort study using longitudinal EHR data from approximately 3 million patients at Mayo Clinic Platform_Accelerate. Cases were patients with confirmed diagnoses of 10 rare diseases; controls (∼5,200 per disease, shared pool) were matched to cases on age, sex, and admission date, and had no record of any rare disease as defined by Orphanet ICD code exclusions (Table 1). The 10 diseases spanned connective tissue disorders (Ehlers-Danlos syndrome classic [EDS-C] and vascular (EDS-V), Marfan syndrome, Loeys-Dietz syndrome), vascular malformations (hereditary haemorrhagic telangiectasia [HHT], Parkes-Weber syndrome), immune conditions (hereditary angioedema [HAE]), metabolic diseases (Fabry disease, Wilson disease), and developmental syndromes (Noonan syndrome). The diseases were chosen based on age of disease onset with sufficient years of documented EHR history, spanning conditions with highly specific early phenotypes (HHT, HAE) to those with insidious, heterogeneous presentations (EDS-C, Loeys-Dietz), to test the generalizability of temporal modeling across the clinical spectrum of rare disease.

**Table 1.** Cohort characteristics.

| Disease | Cases (N) | Controls (N) | % White | % Non-white | % Female | % Male |
| --- | --- | --- | --- | --- | --- | --- |
| Classic Ehlers-Danlos syndrome (EDS-C) | 104 | 5,207 | 93.3% | 6.7% | 78.6% | 21.4% |
| Vascular Ehlers-Danlos syndrome (EDS-V) | 158 | 5,207 | 93.7% | 6.3% | 66.9% | 33.1% |
| Fabry disease | 408 | 5,206 | 84.8% | 15.2% | 53.2% | 46.8% |
| Hereditary angioedema types 1/2 | 768 | 5,205 | 87.6% | 12.4% | 63.7% | 36.3% |
| Hereditary haemorrhagic telangiectasia | 850 | 5,204 | 94.3% | 5.7% | 60.1% | 39.9% |
| Loeys-Dietz syndrome | 690 | 5,206 | 88.1% | 11.9% | 53.8% | 46.2% |
| Marfan syndrome | 935 | 5,206 | 88.3% | 11.7% | 39.6% | 60.4% |
| Noonan syndrome | 236 | 5,207 | 92.8% | 7.2% | 52.3% | 47.7% |
| Parkes-Weber syndrome | 441 | 5,206 | 91.2% | 8.8% | 52.8% | 47.2% |
| Wilson disease | 750 | 5,202 | 92.4% | 7.6% | 51.9% | 48.1% |

### Feature Extraction

HPO phenotype terms were extracted from clinical notes and laboratory results using a medspaCy NLP pipeline; negated, hypothetical, and family-history mentions were excluded. Laboratory values outside the normal range were mapped to corresponding HPO codes using rule-based conversion — capturing biochemical phenotypes that billing codes frequently miss. For example, a documented sweat chloride value ≥ 60 mmol/L was mapped to HP:0012236 (Elevated sweat chloride), and spirometry results with FEV1 < 80% predicted were mapped to HP:0032342 (Reduced FEV1). ICD-9/10 diagnosis codes were collapsed to three-character level, for example I71 (aortic aneurysm), R04 (haemorrhage from respiratory passages), and E83 (disorders of mineral metabolism), capturing disease-relevant patterns while reducing feature sparsity. Three feature sets were evaluated: HPO only, ICD only, and combined HPO+ICD.

### Temporal Tensor Representation

Rather than summarising a patient’s entire clinical history as a single feature vector, temporal models encode records into quarterly time windows — four bins per year, spanning up to 30 years before the index date. Formally, patient records were encoded as a tensor of shape B×T×F, where B is batch size, T is the number of quarterly time bins, and F is the feature dimension. This representation preserves when each clinical finding was recorded, enabling models to learn trajectory patterns such as the rate of phenotypic accrual, the temporal clustering of findings, and the sequence of clinical observations, rather than merely their presence or count. The static baselines (CatBoost, XGBoost, logistic regression) received time-aggregated feature vectors, collapsing all history into a single snapshot.

### Model Architectures

We developed two temporal models based on GRU and Conformer architectures^16,17^. GRU-Attn combines a Gated Recurrent Unit (GRU) network with 3-way attention multi-pooling: attention-weighted, mean, and max-pooled representations of the hidden state sequence are concatenated before the classification head, as depicted in Figure 1. This architecture is designed for the rare disease problem specifically — where the timing of peak phenotypic signals (attention-weighted), the average trajectory (mean pool), and transient early findings (max pool) each carry complementary diagnostic information of uncertain relative importance. The Conformer-Attn (Conformer) model combines local convolutional feature extraction with global self-attention, capturing both short-term phenotypic clusters such as a burst of findings over 2–3 quarters and long-range dependencies such as a finding at year 1 that becomes diagnostically meaningful only in the context of a finding at year 8. This dual local-global inductive bias is particularly suited to rare disease trajectories where informative signals span decades. CatBoost, XGBoost, and logistic regression served as static baselines. All temporal and static models were trained across two independent repeats; results are reported as means (repeat-level SD ≤ 0.011 for all models).

**Figure 1.**
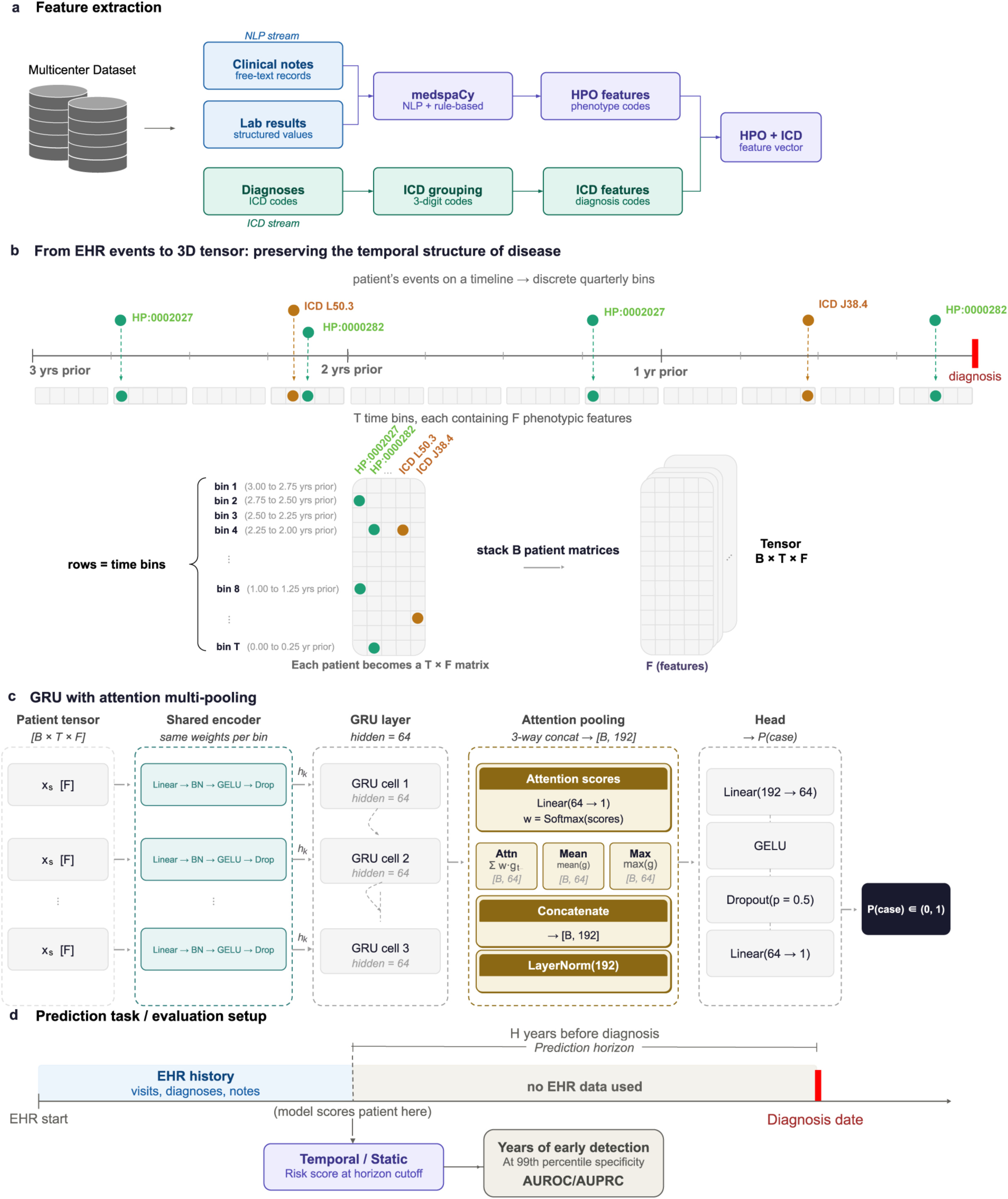
Overview of the longitudinal EHR modeling framework for rare-disease early detection. **a.** Feature extraction pipeline. Clinical notes and laboratory results are processed through an NLP-based stream using medspaCy to extract HPO phenotype features, while structured diagnosis codes are grouped into three-digit ICD features. HPO and ICD features are combined into a unified feature vector. **b.** Construction of the temporal tensor. This example reflects a patient with hereditary angioedema (HAE), where dermatographic urticaria (L50.3), laryngeal edema (J38.4) and multiple occurrences of abdominal pain (HP:0002027) and facial swelling (HP:0000282) are observed. Patient EHR events are aligned relative to the diagnosis date for cases and the last recorded diagnosis date for controls, and divided into discrete quarterly time bins. HPO and ICD features recorded within each bin form a patient-level T × F matrix, and stacking patient matrices creates a B × T × F tensor. This representation preserves when each clinical feature was recorded. **c.** GRU with attention to multi-pooling architecture. Each time bin is encoded using a shared encoder, processed sequentially by a GRU layer, and summarized using attention-weighted, mean, and max pooling before prediction of case probability. **d.** Prediction and evaluation setup. At each prediction horizon, EHR data immediately preceding diagnosis are excluded, and models generate risk scores using only information available before the horizon cutoff. Performance is evaluated using AUROC/AUPRC and early-detection lead time.

### Evaluation

We therefore performed two distinct evaluations. The first was a full-cohort prediction-horizon benchmark using held-out test patients from the complete disease-control cohorts to compare temporal and static models by AUROC and AUPRC. The second was a separate curated diagnostic-odyssey analysis restricted to confirmed prediagnostic cases with extended EHR histories, designed to quantify clinically interpretable lead time before first disease mention and formal diagnosis.

For the full-cohort prediction-horizon benchmark, data were split 60/20/20 (train/validation/test) stratified by case-control status. AUROC and AUPRC were computed at prediction horizons h = 1–10 years by masking the h years immediately preceding diagnosis, so that models at horizon h use only information that is available h years before the patient’s eventual diagnosis.

For the curated diagnostic-odyssey lead-time analysis, early detection was evaluated in a cohort of 143 confirmed prediagnostic cases across 10 diseases, representing 1 to 7% of total cases per disease. This cohort was manually curated using the following criteria: (1) at least 10 years of documented EHR history and (2) expert review to confirm the absence of formal or informal diagnosis before the reference date and the presence of active, undiagnosed symptom burden in clinical notes. This curation enriched for patients with prolonged undiagnosed disease trajectories, in whom phenotypic signals accumulated over years without a unifying diagnosis, rather than patients with incidentally long records. We evaluated two reference periods: Pre-Mention, before first clinical documentation of the target disease, and Pre-Diagnosis, before formal diagnostic code assignment. Detection was defined as a model score exceeding a threshold calibrated to the 99th percentile of the control-score distribution.

## RESULTS

### Cohort

Case counts ranged from 104 (EDS-C) to 935 (Marfan syndrome; Table 1). Controls (∼5,200 per disease, shared pool) were matched to cases on age, sex, and admission date, and had no record of any Orphanet-defined rare disease^18^. The cohort was predominantly White (84.8–94.3%), reflecting the patient population, and was skewed female for connective tissue disorders (EDS-C 78.6% female), consistent with known ascertainment patterns. All disease-control pools were non-overlapping.

### Full-cohort prediction-horizon benchmark

Temporal models outperformed static baselines across all prediction horizons when averaged across 10 diseases, with the advantage widening substantially as the horizon lengthened (Table 2, Figure 2). At h = 1–2 years before diagnosis, GRU-Attn achieved a mean AUROC of 0.928 and Conformer 0.901, compared with 0.871 for CatBoost, the best-performing static model. As the prediction window extended, the performance gap grew: at h = 6–10 years, GRU-Attn (0.726) and Conformer (0.727) maintained meaningful discrimination while CatBoost degraded to 0.693 and, for classic Ehlers-Danlos syndrome specifically, to near chance (0.516). The contrast was most striking for diseases with insidious presentations: for Loeys-Dietz at h = 6–10 years, GRU-Attn (0.722) outperformed CatBoost (0.606) by 0.116 AUROC, a gap that was more negligible at h = 1–2 years (0.930 vs 0.802).

**Figure 2.**
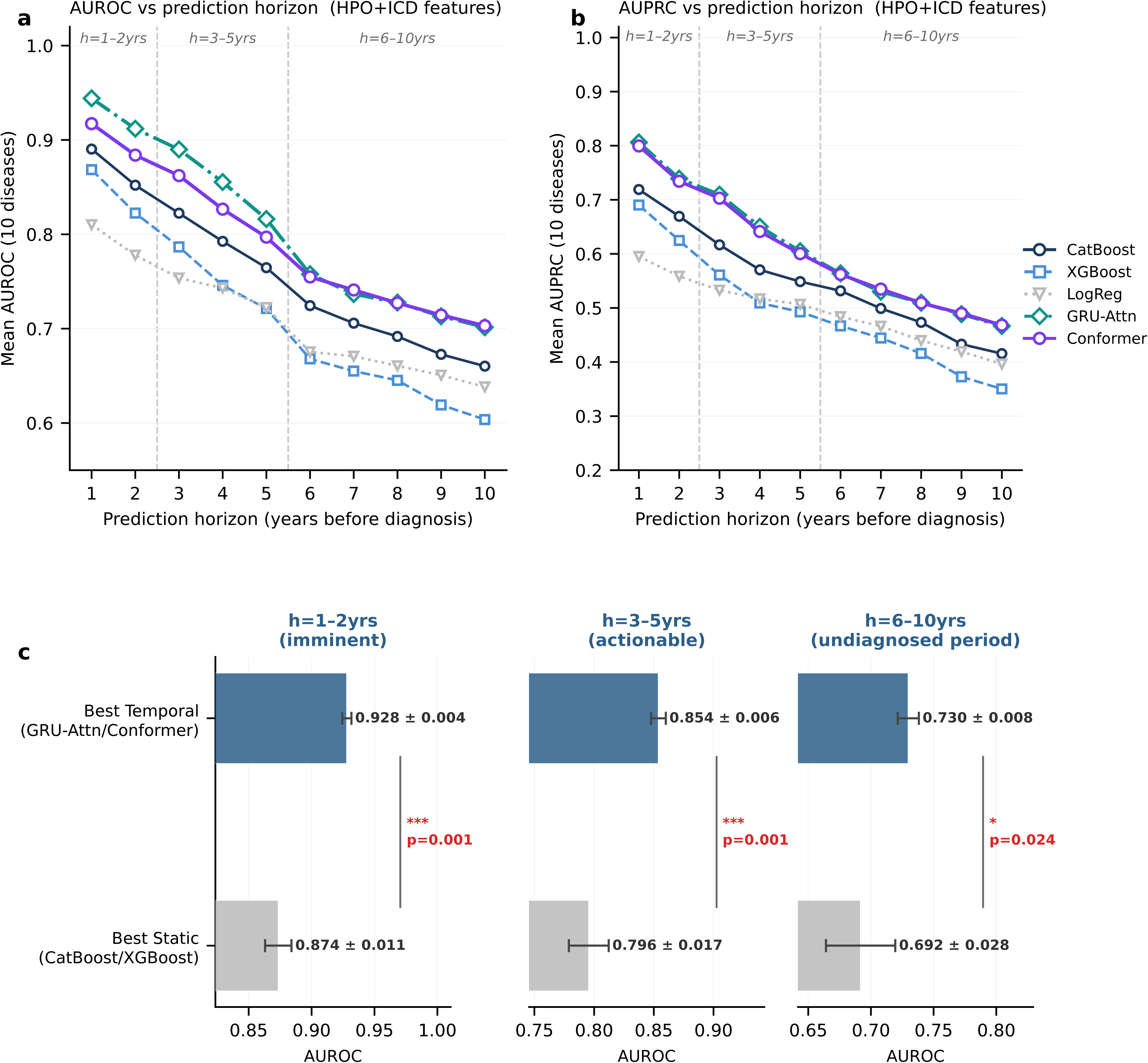
Prediction-horizon model comparison experiment. Model performance was evaluated across fixed prediction horizons using combined HPO and ICD features for 10 rare conditions. At each horizon, information from the years immediately preceding diagnosis was excluded, so model performance reflects discrimination using only data available before that prediction time. **a.** Mean AUROC across prediction horizons for CatBoost, XGBoost, logistic regression, GRU-Attn, and Conformer models. **b.** Mean AUPRC across the same horizons and models. Dashed vertical lines separate the predefined horizon windows: imminent (h = 1–2 years), actionable (h = 3–5 years), and undiagnosed (h = 6–10 years) before diagnosis. **c.** Summary comparison of the best temporal model with the best static baseline within each horizon window. Error bars indicate SEM; p-values compare temporal versus static model performance.

**Table 2.**
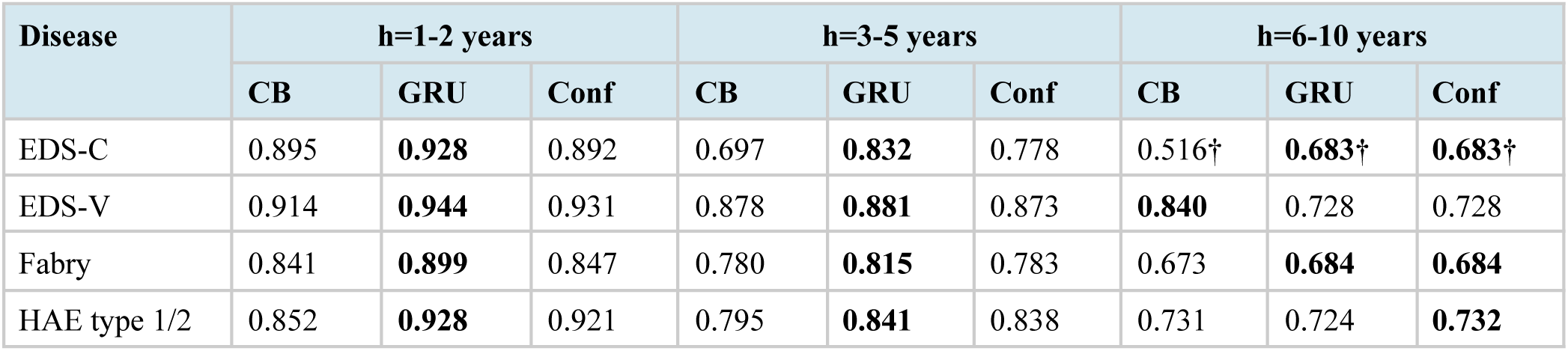
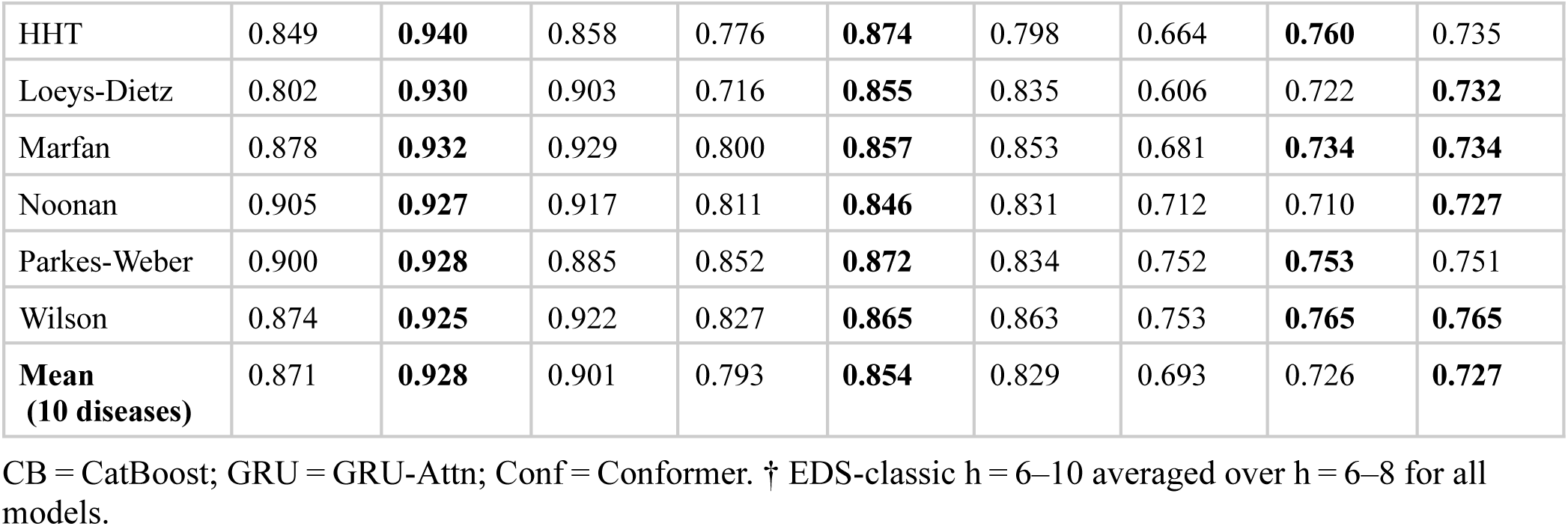
Per-disease AUROC — CatBoost vs GRU-Attn vs Conformer (HPO+ICD, binned horizons). The best-performing model is in bold.

GRU-Attn and Conformer achieved consistently high performance across disease categories and prediction horizon bins. Despite their distinct inductive biases, with GRU-Attn relying on recurrent sequential modeling and Conformer integrating convolutional and self-attention mechanisms, the two architectures yielded comparable results.

### Feature-set Results

Combining HPO and ICD features consistently produced the highest AUROC across all models and horizons (Figure 3). The advantage was largest at short horizons for temporal models: at h = 1–2 years, HPO+ICD improved GRU-Attn AUROC by a mean of +0.063 over the best single source, with positive gains in all 10 diseases (range +0.029 to +0.076). Notably, HPO-only features degraded more slowly than ICD-only features at long horizons; at h = 6–10 years before formal diagnosis the ICD-only AUROC advantage over HPO-only largely disappeared.

**Figure 3.**
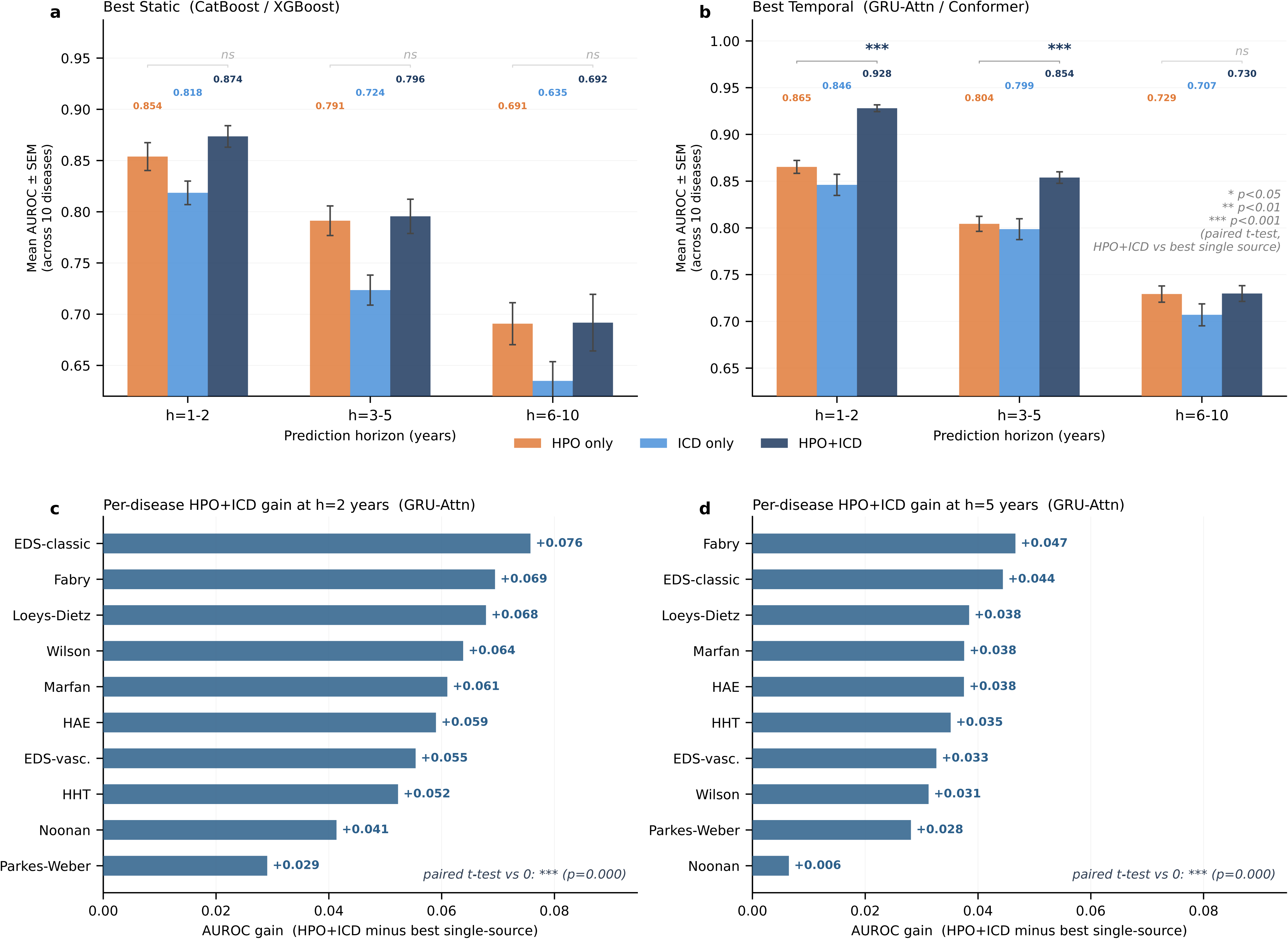
Added value of combining HPO and ICD features in the prediction-horizon benchmark. Model performance was compared using HPO-only, ICD-only, and combined HPO+ICD feature sets across prediction horizons. **a.** Mean AUROC ± SEM for the best static models across the 1–2, 3–5, and 6–10 year prediction windows. **b.** Mean AUROC ± SEM for the best temporal models across the same windows. Combined HPO+ICD features generally achieved the highest performance, particularly for temporal models. **c, d**. Per-disease AUROC gain from combining HPO and ICD features compared with the best single-source feature set at h = 2 years and h = 5 years, respectively.

### Curated diagnostic-odyssey lead-time analysis

The early detection analysis was conducted in a manually curated cohort of 143 confirmed undiagnosed patients across 10 diseases with extended longitudinal EHR history, representing the stratum most consistent with the genuine diagnostic odyssey. Two reference periods were evaluated: Pre-Mention, the most conservative window before any clinical documentation of the target disease, and Pre-Diagnosis, before formal diagnostic code assignment. At 99th-percentile specificity, GRU-Attn identified 96.5% (138/143) cases before first disease mention and 100% (143/143) cases before formal diagnosis (Fig 4a). Detection rates before first disease mention varied in a clinically interpretable pattern: Loeys-Dietz syndrome (80.0%), Marfan syndrome (86%), and vascular Ehlers-Danlos syndrome (88%) were lower, consistent with the more heterogeneous, nonspecific nature of early findings in these conditions; HAE, HHT, and Wilson disease achieved 100%, reflecting earlier and more distinctive phenotypic accrual.

**Figure 4.**
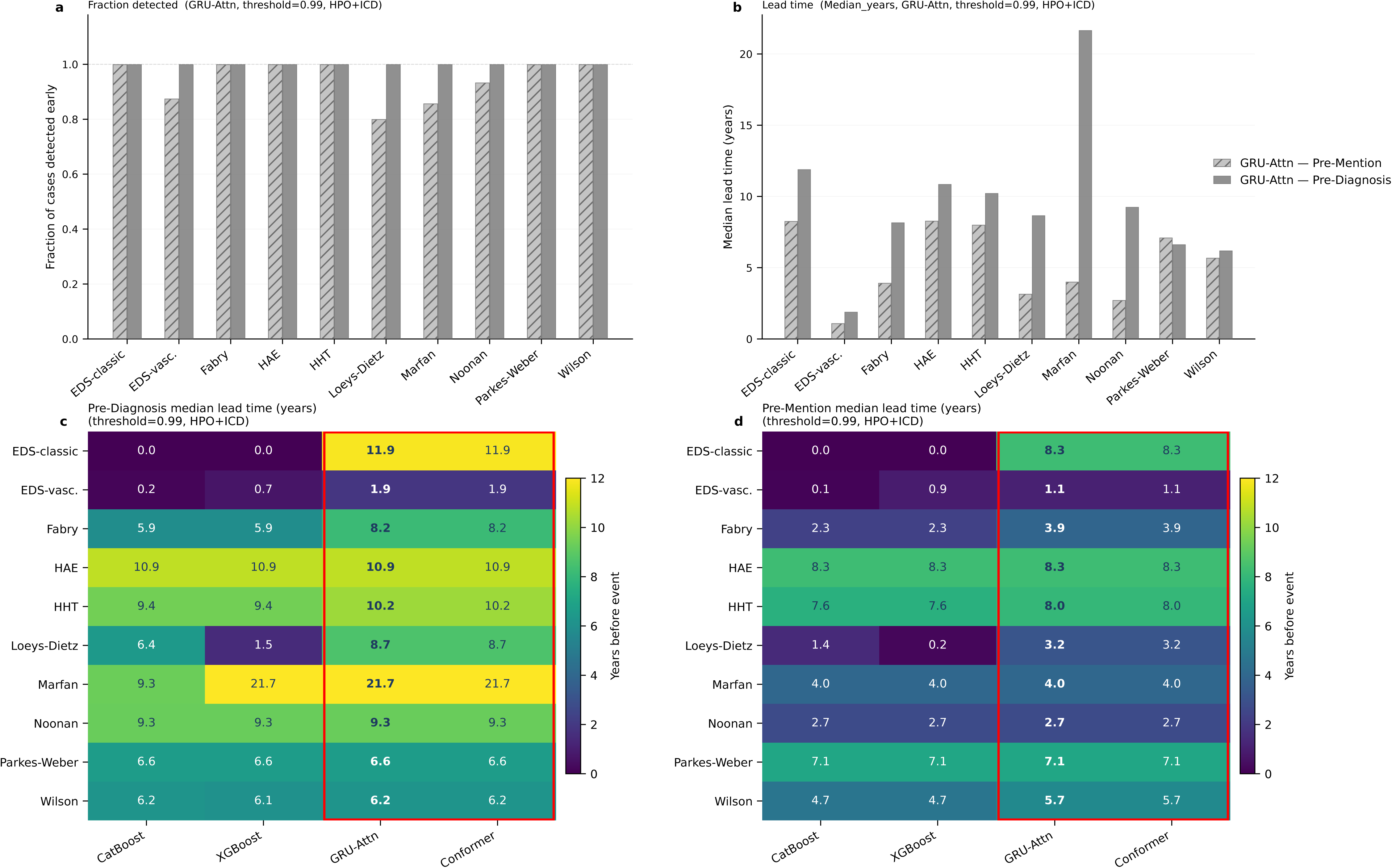
Early detection experiment and lead-time analysis. Early detection was evaluated separately from the prediction-horizon model comparison. For the selected cases with long diagnostic history, model scores were computed using available history before the diagnostic reference date, and a case was considered detected early if its score exceeded the prespecified high-specificity threshold. **a**. Fraction of cases detected before first disease mention and before formal diagnosis using the GRU-Attn model with HPO+ICD features. **b**. Median lead time before first disease mention and before formal diagnosis for each disease. **c**. Median lead time before diagnosis across models and diseases. **d**. Median lead time before first disease mentioned across models and diseases. Red boxes highlight the temporal models, GRU-Attn and Conformer.

At the Pre-Diagnosis reference period, both GRU-Attn and Conformer identified all 143 curated patients before formal diagnosis, with median lead times of 1.9 years for vascular Ehlers-Danlos syndrome to 21.7 years for Marfan syndrome. The 21.7-year median lead time in Marfan syndrome is particularly striking, as it suggests that the relevant phenotypic signal accumulates in the record well before the patient reaches adulthood, opening a window for earlier clinical review, cardiovascular surveillance, and genetic evaluation. Median lead times before first disease mention ranged from 1.1 to 8.3 years across all diseases (Table 3, Figure 4d).

**Table 3.** Early detection performance at 99th-percentile specificity (GRU-Attn, HPO+ICD, Pre-Mention)

| Disease | Prevalence (/100k) | Sensitivity | Specificity | PPV (gen pop) | NNS (gen pop) | PPV (10× enr.) | NNS (10× enr.) |
| --- | --- | --- | --- | --- | --- | --- | --- |
| EDS-C | 2.00 | 100.0% | 99.0% | 0.20% | 501 | 1.96% | 51 |
| EDS-V | 0.40 | 88% | 99.0% | 0.04% | 2858 | 0.35% | 287 |
| Fabry | 2.00 | 100.0% | 99.0% | 0.20% | 501 | 1.96% | 51 |
| HAE type 1/2 | 1.22 | 100.0% | 99.0% | 0.12% | 821 | 1.21% | 83 |
| HHT | 10.00 | 100.0% | 99.0% | 0.99% | 101 | 9.10% | 11 |
| Loeys-Dietz | 0.50 | 80.0% | 99.0% | 0.04% | 2501 | 0.40% | 251 |
| Marfan | 6.50 | 86% | 99.0% | 0.56% | 180 | 5.30% | 19 |
| Noonan | 50.00 | 93% | 99.0% | 4.45% | 22 | 31.86% | 3 |
| Parkes-Weber | 0.30 | 100% | 99.0% | 0.03% | 3334 | 0.30% | 334 |
| Wilson | 3.30 | 100% | 99.0% | 0.33% | 304 | 3.20% | 31 |

\* 10× enriched denotes a specialist-clinic setting with 10-fold higher assumed disease prevalence. Gen pop denotes the general population. PPV and NNS were calculated using published prevalence estimates or representative rare-disease prevalence assumptions where population-based estimates were unavailable (EDS-C and EDS-V<sup>19</sup>, Fabry<sup>20</sup>, HAE<sup>21</sup>, HHT<sup>22</sup>, Loeys-Dietz<sup>23</sup>, Marfan<sup>24</sup>, Noonan<sup>25</sup>, Parkes-Weber<sup>26</sup>, Wilson<sup>27</sup>) and fixed 99.0% specificity; they do not reflect disease prevalence in the curated cohort.

Temporal models demonstrated superior performance on both detection rate and lead time across most diseases (Figure 4c, 4d). GRU-Attn identified all 143 curated cases before formal diagnosis, with median lead times of 21.7 years (Marfan syndrome), 10.9 years (HAE), 10.2 years (HHT), and 8.2 years (Fabry disease).

We evaluated model performance in a simulated clinical deployment setting by computing Positive predictive value (PPV) and Number needed to screen (NNS) using published disease prevalence estimates (Table 3). At 99% specificity, NNS in the general population ranged widely across diseases, from 22 (Noonan) to 3,334 (Parkes-Weber), reflecting the low prevalence of rare diseases. In a 10-fold enriched specialist-clinic setting, NNS improved substantially for higher-prevalence diseases, reaching 3 for Noonan syndrome and 11 for HHT, while remaining above 250 for the three rarest diseases (EDS-V, Loeys-Dietz, and Parkes-Weber; prevalence <1 per 100,000).

### Feature Attribution

SHAP analysis provided face validity by highlighting clinically recognizable disease signatures (Figure 5). For HHT, the top features were telangiectasia and epistaxis, cardinal manifestations appearing years before vascular malformations become symptomatic enough to prompt diagnosis. For HAE, angioedema and abnormal lymphocyte counts dominated, reflecting immune dysregulation detectable in the clinical record long before the specific diagnosis. For Marfan syndrome, aortic aneurysm, aortic root dilation, and pectus excavatum clustered as the top features, findings that individually prompt specialist referral but rarely trigger genetic workup until they co-occur in a recognizable pattern. For EDS-C, congenital musculoskeletal codes (ICD Q79) and joint hypermobility led attribution, reflecting the structural phenotype that accumulates through orthopaedic encounters across childhood and adolescence.

**Figure 5.**
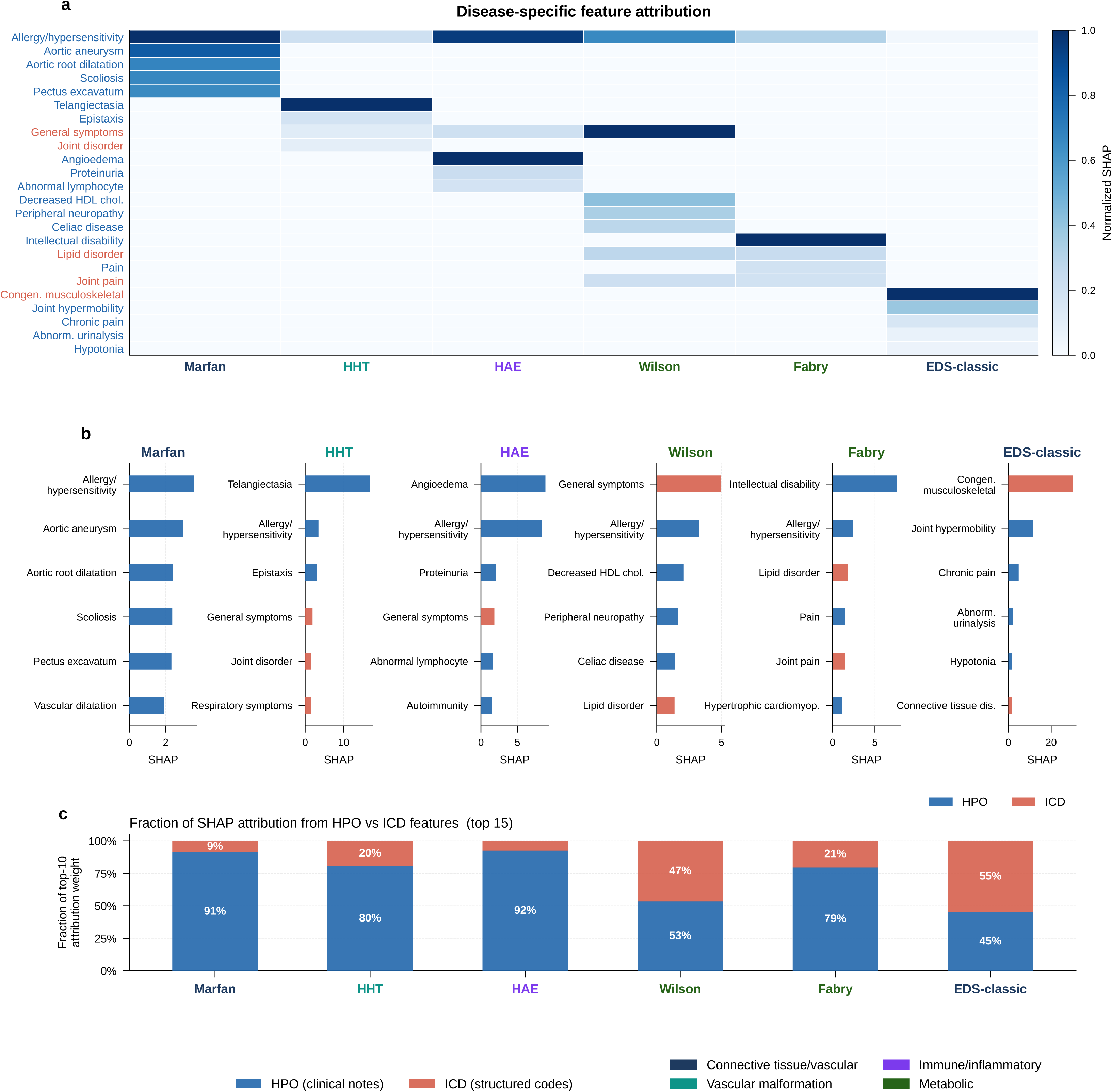
Disease-specific feature attribution. Disease-specific feature attribution was used to identify the clinical features contributing most strongly to model predictions. **a**. Heatmap of normalized attribution values for selected HPO and ICD features across representative rare diseases. **b**. Top contributing features for each disease, colored by feature source: HPO-derived phenotypes from clinical notes and ICD-derived structured diagnosis codes. **c**. Fraction of total attribution contributed by HPO versus ICD features among top-ranked predictors. HPO-derived phenotypes contributed strongly across most diseases, while ICD features provided complementary structured-code signals in selected conditions.

HPO-derived phenotypes from clinical notes contributed the dominant share of attribution weight in most diseases, 80–92% for Marfan, HAE, and HHT, confirming that free-text documentation captures the pre-diagnostic signal more completely than billing codes. Wilson disease and EDS-C were exceptions, with ICD features contributing 47% and 55%, respectively, consistent with Wilson’s reliance on biochemical codes and EDS-C’s structural diagnosis pattern.

## DISCUSSION

Patients with rare diseases accumulate phenotypic signals in the EHR for years, sometimes decades, before a clinician recognizes the pattern. This study demonstrates that temporal deep learning models can recognize those patterns, identifying patients a median of 1.9–21.7 years before formal diagnosis across 10 diverse rare diseases. Prior work has established that machine learning can detect rare diseases early. Our core finding goes further and demonstrates that the temporal trajectory of phenotypic signals carries substantial information beyond their static aggregate; the phenotypic signal strengthens as a patient converges on a diagnosis and this information is potentially actionable in high-specificity retrospective screening simulations, warranting prospective evaluation.

The mechanistic interpretation of the widening performance gap at longer horizons is straightforward. At the time of diagnosis, accumulated phenotypic burden is sufficient for even static models to discriminate. Years earlier, the signal is sparser and its significance depends on temporal context. A patient with Loeys-Dietz syndrome may present with mild aortic root dilation at age 14, a joint hypermobility complaint at 18, and an incidental cervical artery finding at 23, each unremarkable in isolation, each managed by a different specialist, none triggering a unifying diagnosis. The temporal pattern, the rate of accrual and the co-occurrence across time (i.e. the trajectory) is the signal. Static models that collapse this history into a feature count see the same features, but don’t take advantage of when and in what order they appeared. This explains the large temporal advantage for Loeys-Dietz (ΔAUROC +0.139 at h = 3–5) and classic EDS (+0.167 at h = 6–8). For Parkes-Weber and vascular EDS, static models matched or exceeded temporal models at long horizons, confirming the converse: when the early signal is distinctive enough to be detectable from feature presence alone, temporal context adds little. The diseases where temporal modeling matters most are those where the diagnostic odyssey is longest.

Not all diseases benefited equally from temporal modeling, and understanding why is instructive. Vascular EDS and Parkes-Weber syndrome showed the smallest temporal advantage, with static models matching or exceeding GRU-Attn at long horizons. Both conditions share a clinical characteristic that explains this pattern: their early phenotypic presentations are sufficiently distinctive that feature presence alone carries strong discriminative signals. Arterial tortuosity and translucent skin in vascular EDS, and cutaneous vascular malformations in Parkes-Weber, are relatively specific findings that accumulate in the record without requiring temporal context to be informative. By contrast, Loeys-Dietz syndrome and classic EDS show the largest temporal advantage precisely because their early findings, including scattered joint complaints, incidental cardiovascular observations, and nonspecific connective tissue features, are individually unremarkable and only become diagnostically meaningful in the context of their temporal accumulation. The implication for deployment is practical: temporal modeling should be prioritised for conditions with insidious, heterogeneous presentations, while simpler static models may suffice for conditions with early, distinctive phenotypic signatures.

The convergence of GRU-Attn and Conformer across all diseases and horizons is a notable finding in its own right. Two architectures with fundamentally different inductive biases, sequential recurrent processing versus local-global convolutional attention, both produce strong results. This convergence suggests that the temporal representation captures disease-relevant structure that is robust across model classes. Practically, this is reassuring: the benefit of temporal modeling is robust and reproducible, not fully dependent on architectural fine-tuning. For deployment, the simpler GRU-Attn architecture may be preferable given lower computational requirements despite Conformer’s marginal advantage at h ≥ 8.

We demonstrate that combining HPO and ICD features substantially improves over either source alone, and that temporal modeling advances performance further in diseases where the pre-diagnostic trajectory carries information beyond phenotypic burden. The slower degradation of HPO-only features at longer horizons is consistent with billing codes being generated closer to the time of clinical suspicion, while NLP-derived phenotypes capture earlier, uncoded observations in clinical notes. For Parkes-Weber and Noonan syndrome, lead times of static models were equivalent to temporal models, suggesting that specific early phenotypes in these conditions are captured adequately by aggregate features or burden-based scoring. For Marfan syndrome, HAE, HHT, and Fabry disease, temporal models provided 1.2–4.6 additional years of lead time and maintained full detection where static models missed a subset of cases.

Prospective evaluation of this model that includes temporal EHR risk stratification to identify patients for expert review, followed by genetic confirmation for those above a prespecified risk threshold is warranted. This mirrors established paradigms in hereditary cancer surveillance, where population-level EHR risk stratification guides referral to genetic counseling. The lead times observed here, medians of 4–10 years across most diseases, provide a clinically meaningful window for such a pathway. At general population prevalence, PPV is modest (0.03–4.46%), reflecting the mathematical reality of ultra-rare disease screening. Considered as a panel across all 10 conditions, the model achieves 94.2% sensitivity with a pooled PPV of 0.71% at general population prevalence, equivalent to one actionable finding per 140 patients screened. In specialist clinic settings where effective prevalence is 10-fold higher, PPV improves substantially: Noonan (31.9%), HHT (9.1%), and Marfan (5.3%) reach levels comparable to or exceeding accepted cancer screening programs, such as low-dose CT lung cancer screening (PPV ∼3.8%) and multitarget stool DNA testing for colorectal cancer (PPV ∼5%)^28,29^. At 10-fold enrichment, the panel-level PPV reaches 6.70% (NNS 15). For the rarest diseases, including vascular EDS, Loeys-Dietz, and Parkes-Weber, NNS remains above 250 even in enriched settings, suggesting that deployment for these conditions may require additional phenotypic pre-filtering or integration with family history data to achieve actionable yield. Integration via CDS Hooks or SMART on FHIR interfaces would allow deployment within existing EHR workflows without clinical disruption. Alternatively, for patients with prior exome or genome sequencing that did not yield a diagnosis, the model’s sensitivity to an accelerating or new phenotypic trajectory could be used to trigger reanalysis of sequencing data. Both approaches require active EHR monitoring.

The SHAP attribution analysis provided additional face validity by highlighting disease-canonical features, including telangiectasia for HHT, angioedema for HAE, and aortic features for Marfan syndrome, supporting clinician trust in model predictions. The differential reliance on HPO versus ICD features across diseases translates directly into deployment guidance: for HPO-dominant diseases, investment in NLP-based phenotype extraction yields the greatest performance return; for ICD-dominant conditions like Wilson disease and Fabry disease, structured code completeness is equally important.

Limitations include the single-institution Mayo Clinic setting, which may inflate HPO feature richness through comprehensive documentation practices and skew case severity toward complex referral presentations. Subgroup analyses by sex, age, race, ethnicity, and care setting were not performed due to limited case counts per subgroup; evaluating model performance across demographic subgroups and clinical settings remains an important direction for future work. The cohort’s demographic composition may not reflect the broader population of patients with these conditions, and the available data did not support stratified performance analysis by age or sex; such subgroup effects, if present, are not captured here. The use of artificial controls matched for a subset of factors and cases at a higher prevalence than the real world for all 10 diseases. PPV and NNS estimates in Table 3 were therefore calculated using published population prevalence rather than the study case-control ratio. The early detection analysis is restricted to a manually curated subset representing 1–7% of total cases per disease with ≥10 years of confirmed-undiagnosed EHR history, the stratum most amenable to longitudinal monitoring. Analyses were conducted within the Mayo Clinic Platform_Accelerate secure enclave, which imposes access restrictions on the underlying dataset. Prospective validation in an independent, prospectively enrolled cohort across multiple health systems is desirable before clinical deployment.

## CONCLUSION

For millions of patients living through a rare disease diagnostic odyssey, the information needed to make a diagnosis may already be in their medical record, accumulated across years of clinical encounters, but not yet recognized. This study showed that temporal deep learning models can recognize these patterns, identifying patients across 10 rare diseases from 1.9 years before formal diagnosis in vascular Ehlers-Danlos syndrome to 21.7 years in Marfan syndrome. Critically, the advantage widens as the prediction horizon lengthens: static models degrade toward chance years before diagnosis, while temporal models maintain meaningful discrimination. This is most powerful for diseases where the pre-diagnostic trajectory, not its static aggregate, carries the signal. For conditions like Loeys-Dietz syndrome and classic Ehlers-Danlos syndrome, where scattered findings accumulate across a decade of unremarkable encounters, this temporal structure is precisely what enables early detection. By learning from the trajectory of phenotypic signals, temporal models substantially outperform existing approaches, most powerfully for the diseases with heterogeneous pre-diagnostic presentations, where the diagnostic odyssey is longest and the clinical need is greatest. Together, these findings show that the temporal trajectory of EHR-derived phenotypic signals carries independent rare-disease information beyond static phenotypic burden and merits prospective validation as a strategy for rare-disease risk stratification.

## Contributors

L. Yang conceived the study, led all experiments, model development, and analyses, and drafted the manuscript. P. Ng conducted experiments and analyses and assisted in writing the manuscript. M.Y. Lun performed data processing. L. Meyers selected the disease cohort and helped prepare the manuscript. M. Osundiji, as the Mayo Clinic collaborator, provided expertise in rare disease diagnosis and assisted in manuscript preparation. K. Im and A. Kumar reviewed the manuscript and contributed clinical expertise. A. Lavertu contributed to study design and supervised the work. M. Rabinowitz supervised the study.

## Data Sharing Statement

This study involves analysis of de-identified data via the Mayo Clinic Platform_Discover. In accordance with the Code of Federal Regulations, 45 CFR 46.102, the noted activity does not require IRB review. Data shown and reported in this manuscript has been extracted from this environment using an established protocol for data extraction, aimed at preserving patient privacy. The data has been de-identified pursuant to an expert determination in accordance with the HIPAA Privacy Rule. Any data beyond what is reported in the manuscript, including but not limited to the raw EHR data, cannot be shared or released due to the parameters of the expert determination to maintain the data de-identification.

## Acknowledgements

We would like to thank Adam Resnick and Nasibeh Zanjirani Farahani for their helpful advice on accessing the Mayo database.

## One-sentence description

Temporal EHR models trained on longitudinal electronic health records identify rare disease patients with lead times ranging from 1.9 to 21.7 years (median 9.0 years) before formal diagnosis across 10 diseases. These models outperform static machine learning, most substantially for diseases where the pre-diagnostic trajectory, not its aggregate, carries the signal.

## Notes

### Competing Interest Statement

The authors have declared no competing interest.

