## Supplementary Materials for "Temporal EHR Models Detect Rare Disease Years Before Diagnosis"

***Temporal Longitudinal EHR Models for Early Detection of Rare Diseases***

#### Table of Contents

Supplementary Methods

S1. medspaCy HPO Extraction Pipeline

S2. Laboratory-to-HPO Mapping Rules

S3. Model Hyperparameters and Training Details

S4. Statistical Methods

S5. Control cohort

Supplementary Tables

Table S1. Per-disease AUROC at individual prediction horizons h = 1–10 (CatBoost, XGBoost, LogReg, GRU-Attn, Conformer; HPO+ICD)

Table S2. Per-disease AUPRC — CatBoost, XGBoost, LogReg, GRU-Attn, and Conformer (HPO+ICD, binned horizons)

Table S3. Feature-set AUROC by disease and model (HPO-only, ICD-only, HPO+ICD)

Table S4. Early detection performance at 99th-percentile specificity (GRU-Attn, HPO+ICD, Pre-Diagnosis)

Table S5. Laboratory-value-to-HPO mapping rules applied in feature extraction

Supplementary Figures

Figure S1. Early detection before first relevant laboratory investigation (Pre-Lab)

Figure S2. Architecture of the Conformer temporal model.

### Supplementary Methods

#### S1. medspaCy HPO Extraction Pipeline

HPO phenotype terms were extracted from unstructured clinical notes and laboratory reports using a customised medspaCy natural language processing pipeline. The pipeline comprised four sequential stages: (1) sentence segmentation using medspaCy's built-in sentencizer, (2) named entity recognition using a rule-based EntityRuler seeded with HPO term synonyms and abbreviations drawn from the HPO ontology (version 2023-10-09), (3) negation and speculation detection using NegEx rules adapted for clinical English, and (4) context classification to exclude family-history and hypothetical mentions.

Negated terms, hypothetical terms, and terms attributed to family members were excluded from the feature vector. Only terms with an assertion status of 'present' and a subject status of 'patient' were retained. The pipeline was applied to all available free-text notes for each patient, including clinic notes, discharge summaries, radiology reports, and cardiology consultations.

Entity normalisation mapped extracted surface forms to canonical HPO identifiers. Where multiple synonyms mapped to the same HPO code within a single quarterly bin, the feature was recorded as present once (binary encoding). No term frequency weighting was applied within bins.

#### S2. Laboratory-to-HPO Mapping Rules

Laboratory values outside the reference range were systematically mapped to corresponding HPO codes using a curated rule set. Rules were defined as threshold comparisons applied to structured laboratory fields extracted from the EHR. The full mapping table is provided in Table S5. Rules were developed by a clinical expert and applied uniformly across all patient records.

Laboratory-derived HPO features were combined with NLP-extracted HPO features into a single feature vector per quarterly time bin. Where a laboratory value satisfied multiple thresholds, the most specific HPO code was assigned.

#### S3. Model Hyperparameters and Training Details

All models were implemented in PyTorch (version 2.1) and trained on NVIDIA A100 GPUs within the Mayo Clinic Platform_Accelerate secure enclave. Training used the Adam optimiser with a cosine annealing learning rate schedule. Case-control imbalance was addressed using positive class weighting (BCEWithLogitsLoss with pos_weight set to the control-to-case ratio per disease).

GRU-Attn hyperparameters: hidden dimension 256, 2 GRU layers, dropout 0.3, attention heads 4, learning rate $1\times10^{-3}$, batch size 64, maximum epochs 100 with early stopping (patience 10) on validation AUROC.

Conformer hyperparameters: model dimension 128, 4 attention heads, feedforward dimension 512, convolutional kernel size 15, dropout 0.1, learning rate $5\times10^{-4}$, batch size 64, maximum epochs 100 with early stopping (patience 10) on validation AUROC.

CatBoost hyperparameters: depth 6, learning rate 0.05, iterations 500, L2 regularisation 3, class weights proportional to control-to-case ratio.

XGBoost hyperparameters: max depth 6, eta 0.05, n_estimators 500, scale_pos_weight set to control-to-case ratio.

All models were trained across two independent random seeds with fixed 60/20/20 patient-level train/validation/test splits stratified by case-control status. Final results are reported as means across two repeats; repeat-level standard deviation was $\leq 0.011$ for all models and diseases.

#### S4. Statistical Methods

AUROC and AUPRC were computed using scikit-learn (version 1.3). Prediction horizons were implemented by masking all EHR data recorded within h years of the diagnosis date. For binned horizon summaries, AUROC values were averaged across individual horizons within each bin (h = 1–2, h = 3–5, h = 6–10 years). For EDS-C, model data were unavailable beyond h = 8 years; the h = 6–10 bin for EDS-Creflects an average over h = 6–8 for all models.

For the early detection analysis, the specificity threshold was calibrated independently per disease to the 99th percentile of control model scores on the held-out test set. Median lead times were computed as the median time between the model's earliest high-confidence prediction and the reference event across detected cases. No correction for multiple comparisons was applied, as all analyses were pre-specified.

#### S5. Control cohort

Control individuals were selected for each case using stringent matching criteria designed to minimize confounding by age, sex, and healthcare utilization. For every case, eligible controls were drawn from the source population and were required to have a birthdate within ±1 year of the case's birthdate and an identical recorded sex. Controls were further required to have had at least one clinical visit within ±2 months of the case's index date, defined as the date of first disease mention, ensuring that they were actively engaged with the health system over a comparable interval and were therefore subject to similar opportunities for clinical documentation and ascertainment. Finally, controls were required to have no ICD codes corresponding to any rare disease catalogued in the Mondo Disease Ontology rare-disease list (https://mondo.monarchinitiative.org/pages/rare-disease/); any individual carrying a qualifying rare-disease code at any point in their record was excluded.

#### Supplementary Tables

##### Table S1. Per-disease AUROC at individual prediction horizons h = 1–10 (CatBoost, XGBoost, LogReg, GRU-Attn, Conformer; HPO+ICD)

*AUROC values for CatBoost, GRU-Attn, and Conformer at each individual prediction horizon h = 1–10 years before diagnosis, using combined HPO+ICD features. Values are means across two independent training repeats.*

| Disease | h (yrs) | CatBoost | XGBoost | LogReg | GRU-Attn | Conformer |
|---|---|---|---|---|---|---|
| EDS-C | 1 | 0.946 ± 0.00018 | 0.867 ± 0.00012 | 0.619 ± 0.019 | 0.946 ± 0.018 | 0.924 ± 0.032 |
| EDS-C | 2 | 0.844 ± 0.000035 | 0.709 ± 0.000027 | 0.604 ± 0.062 | 0.909 ± 0.00012 | 0.861 ± 0.045 |
| EDS-C | 3 | 0.775 ± 0.00029 | 0.590 ± 0.00017 | 0.571 ± 0.037 | 0.896 ± 0.00025 | 0.835 ± 0.0035 |
| EDS-C | 4 | 0.657 ± 0.000015 | 0.490 ± 0.000091 | 0.597 ± 0.031 | 0.834 ± 0.000078 | 0.767 ± 0.0051 |
| EDS-C | 5 | 0.660 ± 0.000045 | 0.483 ± 0.000097 | 0.614 ± 0.0084 | 0.766 ± 0.00046 | 0.732 ± 0.019 |
| EDS-C | 6 | 0.571 ± 0.00030 | 0.385 ± 0.000079 | 0.578 ± 0.0036 | 0.715 ± 0.00078 | 0.714 ± 0.00020 |
| EDS-C | 7 | 0.524 ± 0.00013 | 0.313 ± 0.00034 | 0.540 ± 0.014 | 0.667 ± 0.00044 | 0.667 ± 0.00024 |
| EDS-C | 8 | 0.452 ± 0.00027 | 0.336 ± 0.00014 | 0.504 ± 0.035 | 0.667 ± 0.00044 | 0.667 ± 0.00024 |
| EDS-V | 1 | 0.924 ± 0.00017 | 0.929 ± 0.00013 | 0.895 ± 0.016 | 0.959 ± 0.00068 | 0.945 ± 0.011 |
| EDS-V | 2 | 0.904 ± 0.00015 | 0.909 ± 0.00024 | 0.864 ± 0.0024 | 0.930 ± 0.00080 | 0.916 ± 0.012 |
| EDS-V | 3 | 0.883 ± 0.00018 | 0.886 ± 0.00021 | 0.790 ± 0.025 | 0.921 ± 0.00097 | 0.911 ± 0.011 |
| EDS-V | 4 | 0.884 ± 0.00030 | 0.846 ± 0.00024 | 0.787 ± 0.0094 | 0.878 ± 0.00022 | 0.869 ± 0.0099 |
| EDS-V | 5 | 0.867 ± 0.00031 | 0.852 ± 0.00028 | 0.768 ± 0.030 | 0.845 ± 0.000056 | 0.840 ± 0.0078 |
| EDS-V | 6 | 0.879 ± 0.00019 | 0.839 ± 0.000054 | 0.733 ± 0.017 | 0.750 ± 0.000 | 0.750 ± 0.00017 |
| EDS-V | 7 | 0.860 ± 0.000 | 0.839 ± 0.00033 | 0.732 ± 0.016 | 0.734 ± 0.00027 | 0.734 ± 0.00045 |
| EDS-V | 8 | 0.860 ± 0.000095 | 0.779 ± 0.00025 | 0.742 ± 0.0014 | 0.734 ± 0.00027 | 0.734 ± 0.00045 |
| EDS-V | 9 | 0.800 ± 0.00035 | 0.719 ± 0.00022 | 0.721 ± 0.029 | 0.719 ± 0.00018 | 0.719 ± 0.00072 |
| EDS-V | 10 | 0.800 ± 0.00016 | 0.700 ± 0.00026 | 0.711 ± 0.043 | 0.703 ± 0.000088 | 0.703 ± 0.00029 |
| Fabry | 1 | 0.848 ± 0.00016 | 0.860 ± 0.00033 | 0.815 ± 0.014 | 0.918 ± 0.00033 | 0.859 ± 0.046 |
| Fabry | 2 | 0.834 ± 0.00026 | 0.841 ± 0.00022 | 0.772 ± 0.0047 | 0.879 ± 0.000084 | 0.835 ± 0.043 |
| Fabry | 3 | 0.812 ± 0.00028 | 0.829 ± 0.00013 | 0.757 ± 0.019 | 0.855 ± 0.0056 | 0.814 ± 0.037 |
| Fabry | 4 | 0.781 ± 0.00013 | 0.794 ± 0.00019 | 0.717 ± 0.040 | 0.816 ± 0.0063 | 0.785 ± 0.025 |
| Fabry | 5 | 0.746 ± 0.00030 | 0.759 ± 0.00028 | 0.674 ± 0.053 | 0.776 ± 0.00059 | 0.749 ± 0.015 |
| Fabry | 6 | 0.713 ± 0.00030 | 0.719 ± 0.000019 | 0.602 ± 0.067 | 0.719 ± 0.00092 | 0.719 ± 0.00036 |
| Fabry | 7 | 0.700 ± 0.00024 | 0.701 ± 0.00016 | 0.591 ± 0.056 | 0.713 ± 0.00029 | 0.713 ± 0.00029 |
| Fabry | 8 | 0.676 ± 0.00012 | 0.689 ± 0.00021 | 0.572 ± 0.040 | 0.689 ± 0.00062 | 0.689 ± 0.00072 |
| Fabry | 9 | 0.640 ± 0.00035 | 0.652 ± 0.000058 | 0.544 ± 0.027 | 0.652 ± 0.00040 | 0.652 ± 0.0010 |
| Fabry | 10 | 0.634 ± 0.00035 | 0.640 ± 0.00017 | 0.540 ± 0.013 | 0.646 ± 0.00048 | 0.646 ± 0.00095 |
| HAE type 1/2 | 1 | 0.862 ± 0.00024 | 0.865 ± 0.00026 | 0.791 ± 0.011 | 0.944 ± 0.00031 | 0.937 ± 0.0013 |
| HAE type 1/2 | 2 | 0.843 ± 0.00018 | 0.848 ± 0.000089 | 0.768 ± 0.0037 | 0.912 ± 0.00038 | 0.906 ± 0.0033 |

| Disease | h (yrs) | CatBoost | XGBoost | LogReg | GRU-Attn | Conformer |
|---|---|---|---|---|---|---|
| HAE type 1/2 | 3 | 0.810 ± 0.000082 | 0.821 ± 0.00023 | 0.759 ± 0.0099 | 0.876 ± 0.00017 | 0.879 ± 0.0085 |
| HAE type 1/2 | 4 | 0.790 ± 0.00017 | 0.797 ± 0.000080 | 0.756 ± 0.0021 | 0.837 ± 0.0038 | 0.830 ± 0.0025 |
| HAE type 1/2 | 5 | 0.784 ± 0.00034 | 0.789 ± 0.00019 | 0.743 ± 0.010 | 0.809 ± 0.000057 | 0.806 ± 0.0062 |
| HAE type 1/2 | 6 | 0.751 ± 0.000014 | 0.743 ± 0.00032 | 0.703 ± 0.0089 | 0.750 ± 0.0039 | 0.750 ± 0.0036 |
| HAE type 1/2 | 7 | 0.741 ± 0.000072 | 0.728 ± 0.000037 | 0.690 ± 0.018 | 0.730 ± 0.00085 | 0.741 ± 0.0042 |
| HAE type 1/2 | 8 | 0.732 ± 0.000079 | 0.719 ± 0.00028 | 0.698 ± 0.0094 | 0.727 ± 0.00034 | 0.734 ± 0.00054 |
| HAE type 1/2 | 9 | 0.722 ± 0.00019 | 0.702 ± 0.000100 | 0.675 ± 0.0078 | 0.717 ± 0.00089 | 0.723 ± 0.0017 |
| HAE type 1/2 | 10 | 0.707 ± 0.00012 | 0.685 ± 0.00026 | 0.651 ± 0.017 | 0.698 ± 0.00055 | 0.710 ± 0.0018 |
| HHT | 1 | 0.870 ± 0.000060 | 0.863 ± 0.00015 | 0.810 ± 0.023 | 0.950 ± 0.00035 | 0.862 ± 0.087 |
| HHT | 2 | 0.829 ± 0.00024 | 0.817 ± 0.00013 | 0.780 ± 0.041 | 0.929 ± 0.00036 | 0.854 ± 0.073 |
| HHT | 3 | 0.804 ± 0.00015 | 0.777 ± 0.000086 | 0.755 ± 0.038 | 0.898 ± 0.00077 | 0.821 ± 0.071 |
| HHT | 4 | 0.784 ± 0.00019 | 0.756 ± 0.000061 | 0.742 ± 0.042 | 0.876 ± 0.0060 | 0.795 ± 0.066 |
| HHT | 5 | 0.740 ± 0.00035 | 0.714 ± 0.00012 | 0.722 ± 0.033 | 0.847 ± 0.0059 | 0.777 ± 0.041 |
| HHT | 6 | 0.697 ± 0.00016 | 0.656 ± 0.00030 | 0.680 ± 0.052 | 0.787 ± 0.0023 | 0.759 ± 0.0041 |
| HHT | 7 | 0.677 ± 0.00013 | 0.652 ± 0.00017 | 0.686 ± 0.027 | 0.778 ± 0.0024 | 0.750 ± 0.0042 |
| HHT | 8 | 0.654 ± 0.00035 | 0.639 ± 0.000084 | 0.680 ± 0.020 | 0.766 ± 0.0019 | 0.738 ± 0.0040 |
| HHT | 9 | 0.648 ± 0.00013 | 0.626 ± 0.000098 | 0.686 ± 0.010 | 0.740 ± 0.0064 | 0.718 ± 0.0087 |
| HHT | 10 | 0.643 ± 0.00021 | 0.618 ± 0.00014 | 0.688 ± 0.0051 | 0.731 ± 0.0066 | 0.712 ± 0.0044 |
| Loeys-Dietz | 1 | 0.837 ± 0.000013 | 0.722 ± 0.00028 | 0.852 ± 0.051 | 0.947 ± 0.00026 | 0.922 ± 0.020 |
| Loeys-Dietz | 2 | 0.767 ± 0.00035 | 0.648 ± 0.00025 | 0.795 ± 0.066 | 0.912 ± 0.000046 | 0.884 ± 0.019 |
| Loeys-Dietz | 3 | 0.741 ± 0.000069 | 0.615 ± 0.000082 | 0.775 ± 0.028 | 0.888 ± 0.00036 | 0.864 ± 0.017 |
| Loeys-Dietz | 4 | 0.721 ± 0.00031 | 0.539 ± 0.000089 | 0.779 ± 0.022 | 0.858 ± 0.00050 | 0.836 ± 0.015 |
| Loeys-Dietz | 5 | 0.685 ± 0.00027 | 0.495 ± 0.00020 | 0.764 ± 0.0092 | 0.817 ± 0.0037 | 0.806 ± 0.0089 |
| Loeys-Dietz | 6 | 0.629 ± 0.00022 | 0.465 ± 0.00015 | 0.738 ± 0.0053 | 0.754 ± 0.0049 | 0.758 ± 0.0016 |
| Loeys-Dietz | 7 | 0.606 ± 0.00035 | 0.449 ± 0.00016 | 0.738 ± 0.0049 | 0.739 ± 0.0053 | 0.748 ± 0.0065 |
| Loeys-Dietz | 8 | 0.621 ± 0.00027 | 0.479 ± 0.00019 | 0.720 ± 0.019 | 0.718 ± 0.011 | 0.726 ± 0.0017 |
| Loeys-Dietz | 9 | 0.598 ± 0.00022 | 0.419 ± 0.000080 | 0.717 ± 0.015 | 0.710 ± 0.015 | 0.722 ± 0.0020 |
| Loeys-Dietz | 10 | 0.575 ± 0.0000096 | 0.403 ± 0.00028 | 0.699 ± 0.020 | 0.688 ± 0.021 | 0.705 ± 0.0022 |
| Marfan | 1 | 0.899 ± 0.00027 | 0.899 ± 0.00034 | 0.869 ± 0.025 | 0.948 ± 0.00022 | 0.945 ± 0.0059 |
| Marfan | 2 | 0.858 ± 0.00013 | 0.856 ± 0.00035 | 0.830 ± 0.028 | 0.917 ± 0.0017 | 0.912 ± 0.0059 |
| Marfan | 3 | 0.834 ± 0.00029 | 0.832 ± 0.00033 | 0.806 ± 0.033 | 0.891 ± 0.0016 | 0.886 ± 0.0066 |
| Marfan | 4 | 0.806 ± 0.00013 | 0.793 ± 0.00010 | 0.773 ± 0.026 | 0.864 ± 0.0027 | 0.858 ± 0.0020 |
| Marfan | 5 | 0.761 ± 0.00033 | 0.755 ± 0.00021 | 0.739 ± 0.021 | 0.816 ± 0.0020 | 0.814 ± 0.0041 |
| Marfan | 6 | 0.711 ± 0.00031 | 0.689 ± 0.00019 | 0.690 ± 0.023 | 0.768 ± 0.0022 | 0.767 ± 0.0037 |
| Marfan | 7 | 0.702 ± 0.00012 | 0.686 ± 0.00019 | 0.683 ± 0.021 | 0.760 ± 0.0021 | 0.759 ± 0.0038 |
| Marfan | 8 | 0.680 ± 0.000070 | 0.672 ± 0.000078 | 0.675 ± 0.021 | 0.734 ± 0.0018 | 0.733 ± 0.0043 |
| Marfan | 9 | 0.666 ± 0.000017 | 0.658 ± 0.00017 | 0.669 ± 0.013 | 0.707 ± 0.0054 | 0.709 ± 0.0043 |
| Marfan | 10 | 0.644 ± 0.00025 | 0.642 ± 0.00027 | 0.658 ± 0.026 | 0.699 ± 0.0054 | 0.700 ± 0.0036 |
| Noonan | 1 | 0.917 ± 0.000041 | 0.875 ± 0.00021 | 0.773 ± 0.046 | 0.948 ± 0.00038 | 0.944 ± 0.024 |

| Disease | h (yrs) | CatBoost | XGBoost | LogReg | GRU-Attn | Conformer |
|---|---|---|---|---|---|---|
| Noonan | 2 | 0.893 ± 0.00017 | 0.849 ± 0.000039 | 0.743 ± 0.054 | 0.906 ± 0.00017 | 0.891 ± 0.021 |
| Noonan | 3 | 0.850 ± 0.00025 | 0.801 ± 0.000045 | 0.728 ± 0.028 | 0.886 ± 0.00056 | 0.875 ± 0.025 |
| Noonan | 4 | 0.820 ± 0.00012 | 0.765 ± 0.00030 | 0.699 ± 0.042 | 0.843 ± 0.00047 | 0.822 ± 0.015 |
| Noonan | 5 | 0.763 ± 0.00030 | 0.738 ± 0.00025 | 0.667 ± 0.026 | 0.808 ± 0.00036 | 0.797 ± 0.017 |
| Noonan | 6 | 0.749 ± 0.00018 | 0.665 ± 0.00026 | 0.582 ± 0.021 | 0.755 ± 0.000074 | 0.766 ± 0.015 |
| Noonan | 7 | 0.724 ± 0.00016 | 0.677 ± 0.000096 | 0.607 ± 0.020 | 0.713 ± 0.00032 | 0.766 ± 0.015 |
| Noonan | 8 | 0.724 ± 0.00018 | 0.652 ± 0.000015 | 0.593 ± 0.037 | 0.723 ± 0.00011 | 0.734 ± 0.015 |
| Noonan | 9 | 0.686 ± 0.00014 | 0.626 ± 0.000056 | 0.598 ± 0.028 | 0.681 ± 0.00031 | 0.691 ± 0.015 |
| Noonan | 10 | 0.675 ± 0.00024 | 0.613 ± 0.000091 | 0.610 ± 0.028 | 0.681 ± 0.00032 | 0.681 ± 0.00032 |
| Parkes-Weber | 1 | 0.913 ± 0.00029 | 0.920 ± 0.000080 | 0.893 ± 0.011 | 0.944 ± 0.0026 | 0.901 ± 0.034 |
| Parkes-Weber | 2 | 0.887 ± 0.00032 | 0.890 ± 0.00034 | 0.873 ± 0.014 | 0.911 ± 0.0038 | 0.869 ± 0.034 |
| Parkes-Weber | 3 | 0.871 ± 0.00023 | 0.879 ± 0.000048 | 0.854 ± 0.030 | 0.899 ± 0.0042 | 0.852 ± 0.0093 |
| Parkes-Weber | 4 | 0.863 ± 0.00022 | 0.862 ± 0.00033 | 0.839 ± 0.037 | 0.878 ± 0.010 | 0.836 ± 0.0093 |
| Parkes-Weber | 5 | 0.823 ± 0.0000024 | 0.822 ± 0.000086 | 0.799 ± 0.038 | 0.840 ± 0.012 | 0.813 ± 0.013 |
| Parkes-Weber | 6 | 0.773 ± 0.00034 | 0.764 ± 0.00021 | 0.753 ± 0.047 | 0.790 ± 0.024 | 0.784 ± 0.031 |
| Parkes-Weber | 7 | 0.767 ± 0.00016 | 0.758 ± 0.00018 | 0.735 ± 0.048 | 0.761 ± 0.025 | 0.761 ± 0.024 |
| Parkes-Weber | 8 | 0.761 ± 0.00023 | 0.741 ± 0.00031 | 0.723 ± 0.057 | 0.756 ± 0.025 | 0.750 ± 0.031 |
| Parkes-Weber | 9 | 0.738 ± 0.00011 | 0.717 ± 0.00015 | 0.691 ± 0.053 | 0.738 ± 0.016 | 0.739 ± 0.016 |
| Parkes-Weber | 10 | 0.721 ± 0.00023 | 0.688 ± 0.00016 | 0.684 ± 0.036 | 0.721 ± 0.016 | 0.721 ± 0.015 |
| Wilson | 1 | 0.887 ± 0.00030 | 0.886 ± 0.000077 | 0.785 ± 0.032 | 0.937 ± 0.0032 | 0.932 ± 0.0020 |
| Wilson | 2 | 0.862 ± 0.00012 | 0.858 ± 0.00026 | 0.750 ± 0.024 | 0.914 ± 0.0048 | 0.911 ± 0.00013 |
| Wilson | 3 | 0.843 ± 0.00021 | 0.836 ± 0.00028 | 0.741 ± 0.024 | 0.889 ± 0.0057 | 0.887 ± 0.00043 |
| Wilson | 4 | 0.819 ± 0.0000038 | 0.816 ± 0.00032 | 0.740 ± 0.019 | 0.869 ± 0.0069 | 0.867 ± 0.00011 |
| Wilson | 5 | 0.818 ± 0.00032 | 0.806 ± 0.00010 | 0.733 ± 0.020 | 0.838 ± 0.0074 | 0.837 ± 0.00083 |
| Wilson | 6 | 0.771 ± 0.000065 | 0.754 ± 0.000031 | 0.695 ± 0.034 | 0.790 ± 0.0092 | 0.780 ± 0.0049 |
| Wilson | 7 | 0.757 ± 0.00022 | 0.747 ± 0.00022 | 0.706 ± 0.026 | 0.770 ± 0.0092 | 0.770 ± 0.00046 |
| Wilson | 8 | 0.757 ± 0.00025 | 0.747 ± 0.000064 | 0.699 ± 0.0069 | 0.764 ± 0.0090 | 0.767 ± 0.0042 |
| Wilson | 9 | 0.754 ± 0.00017 | 0.737 ± 0.000095 | 0.679 ± 0.012 | 0.754 ± 0.0089 | 0.757 ± 0.0042 |
| Wilson | 10 | 0.727 ± 0.000050 | 0.714 ± 0.000063 | 0.654 ± 0.0094 | 0.747 ± 0.0087 | 0.750 ± 0.0040 |

#### Table S2. Per-disease AUPRC — CatBoost, XGBoost, LogReg, GRU-Attn, and Conformer (HPO+ICD, binned horizons)

*Mean AUPRC across two repeats for each disease and model at binned prediction horizons). CB = CatBoost; GRU = GRU-Attn; Conf = Conformer. † EDS-classic h = 6–10 averaged over h = 6–8 for all models.*

| Disease | 1-2 years horizon | | | | | 3-5 years horizon | | | | | 6-10 years horizon | | | | |
|---|---|---|---|---|---|---|---|---|---|---|---|---|---|---|---|
| | CB | XGB | LR | GRU | Conf | CB | XGB | LR | GRU | Conf | CB | XGB | LR | GRU | Conf |
| EDS-C | 0.670 ± 0.000070 | 0.557 ± 0.00029 | 0.322 ± 0.039 | 0.737 ± 0.017 | 0.746 ± 0.0019 | 0.384 ± 0.00015 | 0.171 ± 0.00013 | 0.268 ± 0.021 | 0.552 ± 0.00016 | 0.535 ± 0.022 | 0.204 ± 0.00019 | 0.046 ± 0.00018 | 0.228 ± 0.033 | 0.378 ± 0.00024 | 0.378 ± 0.00027 |
| EDS-V | 0.810 ± 0.000094 | 0.796 ± 0.00011 | 0.752 ± 0.027 | 0.794 ± 0.00015 | 0.792 ± 0.0013 | 0.688 ± 0.00021 | 0.635 ± 0.000079 | 0.644 ± 0.034 | 0.690 ± 0.00028 | 0.688 ± 0.0020 | 0.679 ± 0.00018 | 0.544 ± 0.00025 | 0.529 ± 0.022 | 0.473 ± 0.00017 | 0.473 ± 0.00017 |
| Fabry | 0.573 ± 0.00029 | 0.595 ± 0.00013 | 0.464 ± 0.041 | 0.662 ± 0.00023 | 0.653 ± 0.0078 | 0.508 ± 0.00024 | 0.527 ± 0.00022 | 0.413 ± 0.064 | 0.549 ± 0.0054 | 0.546 ± 0.0047 | 0.393 ± 0.00021 | 0.409 ± 0.00012 | 0.291 ± 0.060 | 0.414 ± 0.00021 | 0.414 ± 0.00021 |
| HAE type 1/2 | 0.699 ± 0.00020 | 0.695 ± 0.00015 | 0.585 ± 0.010 | 0.793 ± 0.00029 | 0.796 ± 0.00093 | 0.614 ± 0.00023 | 0.615 ± 0.000086 | 0.551 ± 0.0030 | 0.656 ± 0.0013 | 0.663 ± 0.0040 | 0.532 ± 0.000097 | 0.509 ± 0.00018 | 0.497 ± 0.0077 | 0.530 ± 0.0011 | 0.539 ± 0.0020 |
| HHT | 0.674 ± 0.00028 | 0.667 ± 0.00021 | 0.616 ± 0.033 | 0.826 ± 0.00035 | 0.783 ± 0.028 | 0.571 ± 0.00015 | 0.546 ± 0.00016 | 0.556 ± 0.038 | 0.721 ± 0.0053 | 0.680 ± 0.019 | 0.470 ± 0.00024 | 0.443 ± 0.00019 | 0.507 ± 0.022 | 0.593 ± 0.0052 | 0.569 ± 0.0045 |
| Loeys-Dietz | 0.668 ± 0.00011 | 0.568 ± 0.000028 | 0.672 ± 0.100 | 0.784 ± 0.00035 | 0.777 ± 0.0043 | 0.537 ± 0.00027 | 0.369 ± 0.00016 | 0.593 ± 0.036 | 0.670 ± 0.0016 | 0.667 ± 0.0023 | 0.430 ± 0.00020 | 0.248 ± 0.00017 | 0.548 ± 0.016 | 0.516 ± 0.011 | 0.525 ± 0.0016 |
| Marfan | 0.707 ± 0.00025 | 0.703 ± 0.00017 | 0.665 ± 0.038 | 0.809 ± 0.0015 | 0.809 ± 0.0028 | 0.590 ± 0.00016 | 0.583 ± 0.00015 | 0.564 ± 0.052 | 0.690 ± 0.0031 | 0.690 ± 0.0034 | 0.465 ± 0.00025 | 0.459 ± 0.00020 | 0.473 ± 0.031 | 0.550 ± 0.0043 | 0.551 ± 0.0035 |
| Noonan | 0.714 ± 0.00024 | 0.582 ± 0.00029 | 0.465 ± 0.031 | 0.784 ± 0.000026 | 0.787 ± 0.015 | 0.565 ± 0.00028 | 0.447 ± 0.00016 | 0.428 ± 0.018 | 0.626 ± 0.00016 | 0.632 ± 0.011 | 0.439 ± 0.00018 | 0.359 ± 0.00017 | 0.338 ± 0.025 | 0.470 ± 0.00017 | 0.487 ± 0.012 |
| Parkes-Weber | 0.725 ± 0.000055 | 0.723 ± 0.00018 | 0.681 ± 0.024 | 0.762 ± 0.0070 | 0.750 ± 0.0068 | 0.673 ± 0.00023 | 0.671 ± 0.00016 | 0.631 ± 0.051 | 0.696 ± 0.012 | 0.679 ± 0.030 | 0.546 ± 0.00015 | 0.528 ± 0.00024 | 0.507 ± 0.053 | 0.558 ± 0.020 | 0.556 ± 0.023 |
| Wilson | 0.702 ± 0.000042 | 0.689 ± 0.00019 | 0.545 ± 0.025 | 0.775 ± 0.0074 | 0.775 ± 0.0020 | 0.657 ± 0.000096 | 0.644 ± 0.00015 | 0.540 ± 0.032 | 0.699 ± 0.0083 | 0.700 ± 0.0028 | 0.583 ± 0.00017 | 0.567 ± 0.00012 | 0.513 ± 0.026 | 0.594 ± 0.0087 | 0.596 ± 0.0042 |
| **Mean (10 diseases)** | 0.694 ± 0.000 | 0.658 ± 0.000 | 0.577 ± 0.037 | 0.773 ± 0.003 | 0.767 ± 0.007 | 0.579 ± 0.000 | 0.521 ± 0.000 | 0.519 ± 0.035 | 0.655 ± 0.004 | 0.648 ± 0.010 | 0.474 ± 0.000 | 0.411 ± 0.000 | 0.443 ± 0.030 | 0.508 ± 0.005 | 0.509 ± 0.005 |

#### Table S3. Feature-set AUROC by disease and model (HPO-only, ICD-only, HPO+ICD)

*Mean AUROC across two repeats for GRU-Attn and Conformer using HPO-only, ICD-only, and combined HPO+ICD feature sets at binned prediction horizons. † EDS-classic h = 6–10 averaged over h = 6–8 for all models.*

| Disease | Model | 1-2 years horizon | | | 3-5 years horizon | | | 6-10 years horizon | | |
|---|---|---|---|---|---|---|---|---|---|---|
| | | HPO | ICD | HPO+ICD | HPO | ICD | HPO +ICD | HPO | ICD | HPO +ICD |
| EDS-C | GRU-Attn | 0.869 ± 0.00020 | 0.763 ± 0.017 | 0.928 ± 0.0091 | 0.770 ± 0.00017 | 0.743 ± 0.010 | 0.832 ± 0.00026 | 0.683 ± 0.00022 | 0.643 ± 0.00010 | 0.683 ± 0.00056 |
| EDS-C | Conformer | 0.869 ± 0.00022 | 0.773 ± 0.013 | 0.892 ± 0.039 | 0.769 ± 0.00020 | 0.740 ± 0.022 | 0.778 ± 0.0093 | 0.682 ± 0.00025 | 0.643 ± 0.0011 | 0.683 ± 0.00022 |
| EDS-V | GRU-Attn | 0.891 ± 0.00016 | 0.854 ± 0.027 | 0.944 ± 0.00074 | 0.839 ± 0.00026 | 0.769 ± 0.026 | 0.881 ± 0.00042 | 0.725 ± 0.00024 | 0.703 ± 0.0048 | 0.728 ± 0.00016 |

| Disease | Model | 1-2 years horizon | | | 3-5 years horizon | | | 6-10 years horizon | | |
|---|---|---|---|---|---|---|---|---|---|---|
| | | HPO | ICD | HPO+ICD | HPO | ICD | HPO +ICD | HPO | ICD | HPO +ICD |
| EDS-V | Conformer | 0.874 ± 0.021 | 0.868 ± 0.0014 | 0.931 ± 0.011 | 0.811 ± 0.035 | 0.781 ± 0.029 | 0.873 ± 0.0095 | 0.701 ± 0.029 | 0.659 ± 0.013 | 0.728 ± 0.00041 |
| Fabry | GRU-Attn | 0.808 ± 0.00034 | 0.826 ± 0.017 | 0.899 ± 0.00021 | 0.752 ± 0.00053 | 0.759 ± 0.012 | 0.815 ± 0.0042 | 0.685 ± 0.00075 | 0.634 ± 0.010 | 0.684 ± 0.00054 |
| Fabry | Conformer | 0.808 ± 0.00010 | 0.808 ± 0.020 | 0.847 ± 0.045 | 0.752 ± 0.00021 | 0.757 ± 0.015 | 0.783 ± 0.026 | 0.684 ± 0.00024 | 0.648 ± 0.0075 | 0.684 ± 0.00067 |
| HAE type 1/2 | GRU-Attn | 0.871 ± 0.0011 | 0.807 ± 0.0077 | 0.928 ± 0.00035 | 0.795 ± 0.00067 | 0.772 ± 0.0083 | 0.841 ± 0.0013 | 0.732 ± 0.00095 | 0.676 ± 0.015 | 0.724 ± 0.0013 |
| HAE type 1/2 | Conformer | 0.872 ± 0.00017 | 0.801 ± 0.0064 | 0.921 ± 0.0023 | 0.796 ± 0.0015 | 0.782 ± 0.0066 | 0.838 ± 0.0057 | 0.734 ± 0.00062 | 0.712 ± 0.0020 | 0.732 ± 0.0023 |
| HHT | GRU-Attn | 0.883 ± 0.00025 | 0.883 ± 0.012 | 0.940 ± 0.00036 | 0.822 ± 0.0027 | 0.841 ± 0.0075 | 0.874 ± 0.0042 | 0.755 ± 0.0037 | 0.725 ± 0.011 | 0.760 ± 0.0039 |
| HHT | Conformer | 0.885 ± 0.00027 | 0.883 ± 0.0030 | 0.858 ± 0.080 | 0.825 ± 0.0013 | 0.839 ± 0.014 | 0.798 ± 0.059 | 0.759 ± 0.0017 | 0.741 ± 0.0062 | 0.735 ± 0.0051 |
| Loeys-Dietz | GRU-Attn | 0.864 ± 0.000092 | 0.821 ± 0.0016 | 0.930 ± 0.00015 | 0.806 ± 0.00012 | 0.764 ± 0.0098 | 0.855 ± 0.0015 | 0.725 ± 0.00011 | 0.651 ± 0.027 | 0.722 ± 0.011 |
| Loeys-Dietz | Conformer | 0.864 ± 0.000092 | 0.792 ± 0.058 | 0.903 ± 0.019 | 0.806 ± 0.00012 | 0.765 ± 0.045 | 0.835 ± 0.014 | 0.725 ± 0.00011 | 0.697 ± 0.0021 | 0.732 ± 0.0028 |
| Marfan | GRU-Attn | 0.873 ± 0.00027 | 0.852 ± 0.0016 | 0.932 ± 0.00095 | 0.807 ± 0.00016 | 0.808 ± 0.0016 | 0.857 ± 0.0021 | 0.732 ± 0.00028 | 0.699 ± 0.00100 | 0.734 ± 0.0034 |
| Marfan | Conformer | 0.873 ± 0.00028 | 0.857 ± 0.0031 | 0.929 ± 0.0059 | 0.807 ± 0.00018 | 0.811 ± 0.0022 | 0.853 ± 0.0042 | 0.732 ± 0.00020 | 0.712 ± 0.0033 | 0.734 ± 0.0039 |
| Noonan | GRU-Attn | 0.878 ± 0.000086 | 0.879 ± 0.0051 | 0.927 ± 0.00028 | 0.801 ± 0.00022 | 0.837 ± 0.0062 | 0.846 ± 0.00046 | 0.723 ± 0.00024 | 0.700 ± 0.0033 | 0.710 ± 0.00023 |
| Noonan | Conformer | 0.878 ± 0.00011 | 0.841 ± 0.029 | 0.917 ± 0.022 | 0.800 ± 0.0027 | 0.806 ± 0.028 | 0.831 ± 0.019 | 0.723 ± 0.00020 | 0.675 ± 0.039 | 0.727 ± 0.012 |
| Parkes-Weber | GRU-Attn | 0.858 ± 0.00022 | 0.887 ± 0.00049 | 0.928 ± 0.0032 | 0.829 ± 0.00015 | 0.843 ± 0.00039 | 0.872 ± 0.0090 | 0.759 ± 0.00040 | 0.759 ± 0.000077 | 0.753 ± 0.021 |
| Parkes-Weber | Conformer | 0.858 ± 0.00022 | 0.882 ± 0.0021 | 0.885 ± 0.034 | 0.829 ± 0.00015 | 0.842 ± 0.0028 | 0.834 ± 0.011 | 0.759 ± 0.00040 | 0.760 ± 0.0017 | 0.751 ± 0.023 |
| Wilson | GRU-Attn | 0.855 ± 0.00030 | 0.858 ± 0.0025 | 0.925 ± 0.0040 | 0.820 ± 0.00037 | 0.817 ± 0.0023 | 0.865 ± 0.0067 | 0.768 ± 0.00047 | 0.734 ± 0.0053 | 0.765 ± 0.0090 |
| Wilson | Conformer | 0.855 ± 0.000 | 0.854 ± 0.0047 | 0.922 ± 0.0011 | 0.820 ± 0.00024 | 0.825 ± 0.0031 | 0.863 ± 0.00046 | 0.768 ± 0.00042 | 0.751 ± 0.012 | 0.765 ± 0.0036 |

#### Table S4. Early detection performance at 99th-percentile specificity (GRU-Attn, HPO+ICD, Pre-Diagnosis)

*Pre-Diagnosis early detection results for GRU-Attn with HPO+ICD features at 99th-percentile specificity. All 10 diseases achieved 100% detection before formal diagnosis. PPV and NNS calculated using the same published prevalence estimates as Table 3 in the main text. 10× enriched denotes a specialist-clinic setting.*

| Disease | Sensitivity | Specificity | PPV (gen pop) | NNS (gen pop) | PPV (10× enr.) | NNS (10× enr.) | Median lead (yrs) |
|---|---|---|---|---|---|---|---|
| EDS-C | 100.0% | 99.0% | 0.20% | 501 | 1.96% | 51 | 11.9 |
| EDS-V | 100.0% | 99.0% | 0.04% | 2501 | 0.40% | 251 | 1.9 |
| Fabry | 100.0% | 99.0% | 0.20% | 501 | 1.96% | 51 | 8.2 |
| HAE type 1/2 | 100.0% | 99.0% | 0.12% | 821 | 1.21% | 83 | 10.9 |
| HHT | 100.0% | 99.0% | 0.99% | 101 | 9.10% | 11 | 10.2 |
| Loeys-Dietz | 100.0% | 99.0% | 0.05% | 2001 | 0.50% | 201 | 8.7 |
| Marfan | 100.0% | 99.0% | 0.65% | 155 | 6.11% | 16 | 21.7 |
| Noonan | 100.0% | 99.0% | 4.76% | 21 | 33.44% | 3 | 9.3 |
| Parkes-Weber | 100.0% | 99.0% | 0.03% | 3334 | 0.30% | 334 | 6.6 |
| Wilson | 100.0% | 99.0% | 0.33% | 304 | 3.20% | 31 | 6.2 |

#### Table S5. Laboratory-value-to-HPO mapping rules applied in feature extraction

*Example set of rule-based laboratory-to-HPO mappings applied during feature extraction. Threshold values were defined by a clinical expert prior to analysis. ULN = upper limit of normal; F = female; M = male.*

| Laboratory Finding | Rule / Threshold | HPO Code | HPO Term |
|---|---|---|---|
| Sweat chloride | ≥ 60 mmol/L | *HP:0012236* | Elevated sweat chloride |
| Spirometry FEV1 | < 80% predicted | *HP:0032342* | Reduced FEV1 |
| Serum ceruloplasmin | < 20 mg/dL | *HP:0002155* | Hypoceruloplasmineinia |
| 24-hr urine copper | ≥ 100 µg/day | *HP:0003527* | Increased urinary copper |
| Alpha-galactosidase A | < 1.5 nmol/hr/mg (males) | *HP:0003341* | Deficiency of alpha-galactosidase A |
| Lyso-Gb3 | > 2 nmol/L | *HP:0003341* | Deficiency of alpha-galactosidase A |
| C1 inhibitor level | < 50% normal | *HP:0004350* | Abnormality of C1 esterase inhibitor |
| C4 complement | Low (persistently) | *HP:0005421* | Decreased serum complement C4 |
| Hemoglobin | < 12 g/dL (F) / < 13 g/dL (M) | *HP:0001903* | Anemia |
| ALT/AST | > 3× ULN | *HP:0002910* | Elevated hepatic transaminases |
| Creatinine kinase | > 2× ULN | *HP:0003236* | Elevated serum creatine kinase |
| Proteinuria | > 300 mg/day | *HP:0000093* | Proteinuria |
| eGFR | < 60 mL/min/1.73m² | *HP:0012622* | Chronic kidney disease |
| Serum ferritin | < 12 µg/L | *HP:0001824* | Weight loss |

### Supplementary Figures

#### Figure S1. Early detection before first relevant laboratory investigation (Pre-Lab)

Early detection performance evaluated using the Pre-Lab reference period, defined as the period before first relevant laboratory investigation for the target disease. a. Fraction of curated cases detected before first laboratory investigation (GRU-Attn, HPO+ICD, threshold = 99th-percentile specificity). b. Median lead time before first laboratory investigation among detected patients (GRU-Attn, HPO+ICD). c. Heatmap of median lead time before first laboratory investigation among detected patients across all five models (CatBoost, XGBoost, GRU-Attn, Conformer). Cell annotations show median lead time in years with percentage of cases detected early in parentheses. Red box highlights temporal models (GRU-Attn and Conformer).

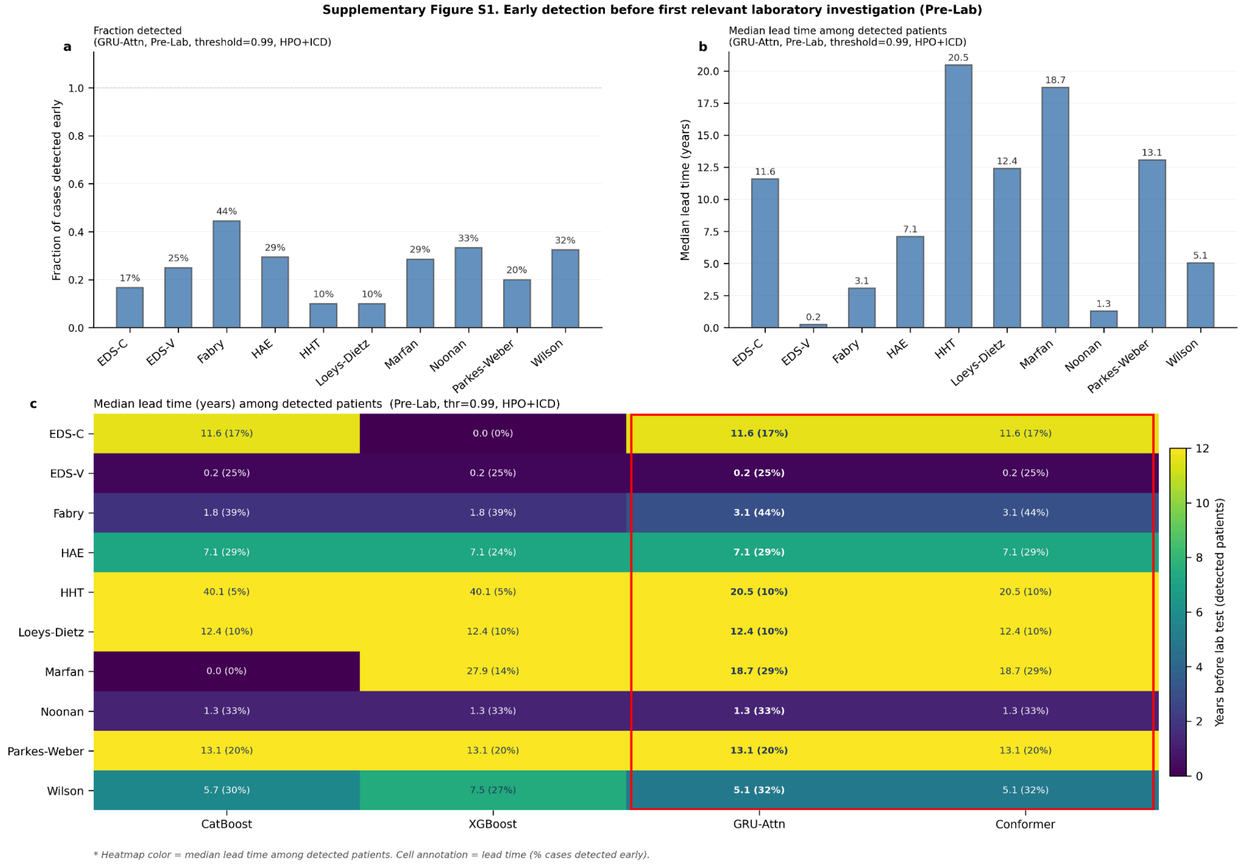


**Figure S2. Architecture of the Conformer temporal model.** Each patient's EHR history is encoded as a quarterly time-binned sequence and passed through a shared feature encoder (MLP with batch normalisation). The encoded sequence is processed by a stack of Conformer blocks, each combining a local depthwise convolutional layer for short-range temporal feature extraction with a multi-head self-attention layer for long-range dependency modelling, interleaved with feed-forward sublayers and residual connections. A learnable classification token prepended to the sequence aggregates global context across all time bins. The final classification token representation is passed through a linear head to produce a case probability score.

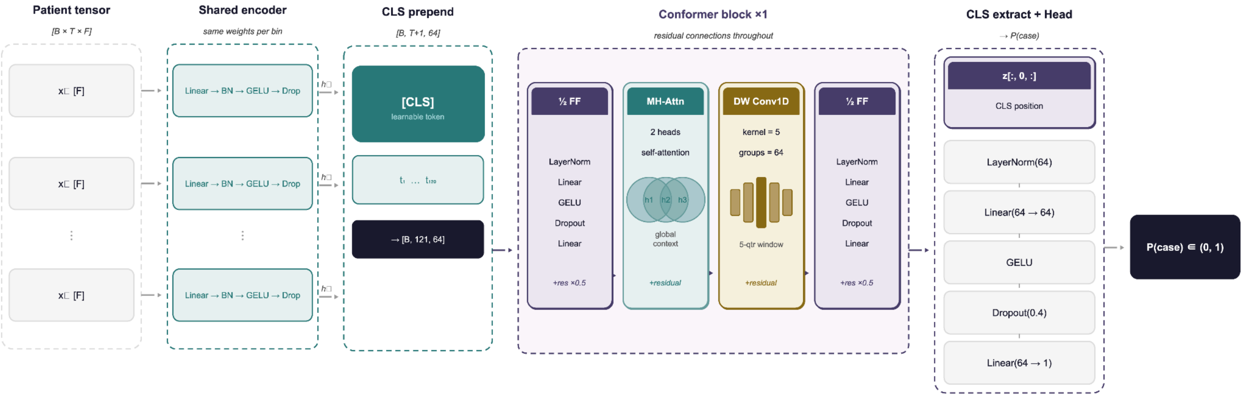
Patient tensor
[B × T × F]
x□ [F]
x□ [F]
x□ [F]
Shared encoder
same weights per bin
Linear → BN → GELU → Drop
Linear → BN → GELU → Drop
Linear → BN → GELU → Drop
h□
h□
h□
CLS prepend
[B, T+1, 64]
[CLS]
learnable token
$t_1$ … $t_{120}$
→ [B, 121, 64]
Conformer block ×1
residual connections throughout
½ FF
LayerNorm
Linear
GELU
Dropout
Linear
+res ×0.5
MH-Attn
2 heads
self-attention
h1
h2
h3
global context
+residual
DW Conv1D
kernel = 5
groups = 64
5-qtr window
+residual
½ FF
LayerNorm
Linear
GELU
Dropout
Linear
+res ×0.5
CLS extract + Head
→ P(case)
z[:, 0, :]
CLS position
LayerNorm(64)
Linear(64 → 64)
GELU
Dropout(0.4)
Linear(64 → 1)
P(case) ∈ (0, 1)
